# Neurometabolite differences in Autism as assessed with Magnetic Resonance Spectroscopy: a systematic review and meta-analysis

**DOI:** 10.1101/2024.02.07.24302277

**Authors:** Alice R. Thomson, Duanghathai Pasanta, Tomoki Arichi, Nicolaas A. Puts

## Abstract

*^1^H-Magnetic Resonance Spectroscopy (MRS)* is a non-invasive technique that can be used to quantify the concentrations of metabolites in the brain in vivo. MRS findings in the context of autism are inconsistent and conflicting. We performed a *systematic review and meta-analysis* of MRS studies observing glutamate and gamma-aminobutyric acid (GABA), as well as brain metabolites involved in energy metabolism (glutamine, creatine), neural integrity (e.g. n-acetyl aspartate (NAA), choline, myo-inositol) and neuro-inflammation (glutathione) in autism cohorts. Data were extracted and grouped by metabolite, brain region and several other factors before calculation of standardised effect sizes. Overall, we find significantly lower concentrations of GABA and NAA in autism, indicative of disruptions to the balance between excitation/inhibition within brain circuits, as well as neural integrity. Further analysis found these alterations are most pronounced in autistic children and in limbic brain regions relevant to autism phenotypes. Additionally, we show how study outcome varies due to demographic and methodological factors (e.g. medication use), emphasising the importance of conforming with consensus study designs and transparent reporting.

**Highlights:**

- We performed a meta-analysis of *Magnetic Resonance Spectroscopy* findings in autism.
- We find significantly lower concentrations of GABA and NAA in children with autism.
- These alterations were most pronounced in limbic brain regions.
- Demographic factors (including sex) contribute to MRS study outcome variation.

## 1. Introduction

### 1.1 background

Autism Spectrum Condition (henceforth referred to as ‘autism’ as preferred by the autistic community) is a neurodevelopmental condition, affecting 1-2% of the population and characterised by differences in cognition, social communication, and repetitive and restricted interests (American Psychiatric Association, 2000, 2013; Horder et al., 2018). Sensory differences are also extremely common and are reported in 60-95% of autistic individuals (He et al., 2021; Klintwall et al., 2011; Rogers & Ozonoff, 2005).

Most cases of autism are idiopathic in nature (Horder et al., 2018) with around a thousand gene variants linked to the condition in humans (Grove et al., 2019). Stratification of patient aetiologies based on shared neurobiological ‘endpoints’ of this genetic heterogeneity is essential to advance our understanding of the neurodevelopmental abnormalities that underpin autism at the biochemical, neuronal and circuit level (Brian et al., 2016; Sanders, 2015; Wolfers et al., 2019) One such common ‘endpoint’ across autism aetiologies might be a disruption in the balance of excitation/inhibition in brain circuits (E/I; Rubenstein & Merzenich, 2003). Typical brain states rely on a homeostasis between E/I, predominantly determined by the endogenous activity of glutamate (Glu) and gamma-aminobutyric acid (GABA), the adult human brain’s principal excitatory and inhibitory neurotransmitters respectively. The maintenance of this E/I balance is a critical component of typical neurodevelopment and neuronal computation (Ghisleni et al., 2015; Rubenstein & Merzenich, 2003; S. Zhou & Yu, 2018).

In autism, an ‘E/I’ imbalance within brain circuits is thought to lead to differences in sensory function, memory, and social and non-social cognition (Rubenstein & Merzenich, 2003). Several lines of evidence support this theory, including genetic, preclinical animal, and post-mortem studies of human brain tissue (Ajram et al., 2019; DeLorey et al., 2008; Etherton et al., 2011; Fatemi et al., 2002; Hashemi et al., 2017; Moreno-De-Luca et al., 2013; Port et al., 2019; Tuchman & Rapin, 2002; Yan et al., 2005). For example, studies have found various autism-associated genes with a role in neuronal excitatory and inhibitory cascades (Grove et al., 2019; Havdahl et al., 2021; Moreno-De-Luca et al., 2013). Other studies have found differences in inhibitory interneuron availability and synaptic pruning (Hashemi et al., 2017; Kim et al., 2017; Tang et al., 2014), and many animal models of autism show differences in E/I signalling while showing autistic phenotypes (DeLorey et al., 2008; Etherton et al., 2011; Qi et al., 2022). Finally, seizure disorders, thought to arise due to abnormally increased excitation brain circuits (Rubenstein & Merzenich, 2003) regularly co-occur with autism (ranging from 5-40%; Tuchman & Rapin, 2002).

However, other biological theories exist; the redox hypothesis is an additional (although not necessarily discrete) aetiological model of autism in which environmentally induced oxidative stress leads to altered neurodevelopment in genetically vulnerable individuals (Endres et al., 2017). Consistent with this, post-mortem tissue samples from autistic individuals show evidence of an abnormal redox state and glutathione (GSH) deficiency (Adams et al., 2009; Chauhan et al., 2012; F. Gu et al., 2013). GSH is a cellular antioxidant which donates electrons for the reduction of reactive oxygen species (ROS e.g hydrogen peroxide) within neurons and glial cells (Dringen, 2000) and thus acts as a crucial defence against neuronal oxidative stress (Dringen, 2000). Preclinical researchers also find reductions in GSH brain concentrations in mouse models of autism, with further GSH depletion increasing the occurrence of autism-like behaviours (Nadeem et al., 2019). (Bjørklund et al., 2020; Frye et al., 2013).

*^1^H-Magnetic Resonance Spectroscopy (MRS)* is a non-invasive technique that can be used to quantify the concentrations of endogenous brain metabolites including GSH, Glu and GABA in the human brain in vivo (Puts & Edden, 2012). As proxy measures of E/I balance (Glu/GABA) and neuroinflammation (GSH), MRS is therefore highly valuable for validating the E/I imbalance/redox hypothesis of autism in humans and so identifying human biochemical pathways for invention.

Beyond GABA/Glu and GSH, other MRS measured metabolites have been implicated in the pathophysiology of autism, including those involved in energy metabolism (glutamine (Gln) and creatine (Cr)), and neural cytoarchitecture and integrity (e.g. N-acetyl aspartate (NAA), choline (Cho), myo-inositol (mI); Levitt et al., 2003; Libero et al., 2016; O’Neill et al., 2020; Oner et al., 2007; Vasconcelos et al., 2008), although preclinical evidence complementing this work is more limited.

### 1.2 MRS

MRS works by suppressing the brain water signal to detect other ^1^H-proton containing metabolites at lower concentrations (in the order of 0 – 20 mM). ^1^H protons in different metabolites have different chemical environments (e.g. surrounded by more electrons than others) giving each metabolite a unique resonant (& spatial) frequency measured in parts per million (ppm). This spatial frequency is plotted along a chemical shift axis. The amplitude of each metabolite peak along this axis is proportional to the number of ^1^H protons detected and as such the metabolite concentration (Puts & Edden, 2012). To obtain an acceptable signal to noise ratio (SNR), metabolites are quantified from an average MRS spectrum, the result of many repeated measurements (transients) from the same voxel.

Conventional MR pulse sequences for MRS acquisition, including Point Resolved Spectroscopy (PRESS; Bottomley, 1987) and Stimulated Echo Acquisition Mode (STEAM; Frahm et al., 1987), can detect and so quantify high-concentration metabolites including NAA, total choline and total creatine. Tailored ‘edited’ MRS sequences are required to resolve GABA, Glu and GSH, as their signals are usually masked by metabolites in greater concentrations (e.g. NAA) and so with stronger resonances (Mullins et al., 2014). One such edited approach is MEshcher-GArwood Point RESolved Spectroscopy sequence (MEGA-PRESS; Mescher et al., 1996, 1998). MEGA-PRESS uses a 1.9 ppm editing pulse to selectively modulate the GABA signal during the acquisition phase (edit-ON). Subtraction of this edited spectra (edit-ON) from an unedited spectrum (edit-OFF) gives the difference (DIFF) spectrum, which contains metabolite frequencies modulated by the GABA-selective editing, including GABA at 3.02ppm. A similar approach can be used for GSH by placing editing-ON pulses at 4.56 ppm. Due to contamination with macromolecules which are also affected by the GABA-selective editing pulse, GABA signal is often denoted as GABA+ (GABA + macromolecules; Mullins et al., 2014). Similarly, due to the poor chemical shift dispersion achieved at 1.5-3 Tesla (overlap of metabolite spatial frequencies on the chemical shift axis) it is often not possible to distinguish Glu signals from that of its precursor glutamine (Gln). Glx therefore describes the combined concentrations of Gln and Glu (Mullins et al., 2014).

### 1.3 The current study

While MRS has previously been used to identify autism-associated alterations in brain metabolite concentrations, variation in brain region, ^1^H-MRS parameters, and small sample sizes have limited interpretation, with highly conflicting outcomes.

Here, we quantitively and systemically review previous MRS work in autism, grouping results by brain region and, as metabolite concentrations have been shown to vary across the lifespan, by age (Ghisleni et al., 2015; Porges et al., 2021; Reyngoudt et al., 2012). To extensively cover the previous literature, we focus on seven key metabolites; GABA, Glu/Glx, Gln, NAA, creatine, choline and mI, these being the most widely studied metabolites in autism (Ford & Crewther, 2016a). We aimed to comprehensively summarise the MRS-based evidence for autism associated alterations in these brain metabolite concentrations and as such, autism aetiological theories. We performed appropriate assessments for MRS data quality, which have not yet been performed in the autism field, and controlled for non-independence of data, as most studies report more than one metabolite and/or measure from more than one brain region per participant. We additionally investigate potential sources of bias within the included studies such as MRS data acquisition parameters, demographic variables, and quality of MRS data. In doing so we aimed to isolate factors that contributed to the heterogenicity of previous findings and as such guide researchers towards more replicable study designs. Finally, we discuss the potential source of metabolite differences in autism at the mechanistic level.

## 2. Materials and methods

### 2.1 Search criteria and study selection strategy

A systematic search of databases (Ovid Medline, Pubmed, Web of Science and Google Scholar) was preformed using the search terms selected using the Boolean method from *litserchr* in R (29/09/22), these being; *(“ASD” OR “Autism Spectrum Disorder” OR “Autistic disorder” OR “Asperger’s Syndrome” OR “Autism”) AND (“1H-MRS” OR “in vivo magnetic resonance spectroscopy” OR “magnetic resonance spectroscopy” OR “proton magnetic resonance spectroscopy” OR “MRS” OR “GABA” OR “Glutamate” OR “Creatine” OR “Choline” OR “metabolite”*). Duplicate papers were removed before abstract screening using the *metagear* package in R (Lajeunesse, 2016). **Papers were included if they met all of the following criteria; (1) use of in-vivo MRS to measure metabolites in the brain; (2) investigation of brain metabolite concentrations in autism and non-autism groups (3) MRS measured from a specified brain region and (4) the study was published in a peer-reviewed journal, and was written in English or translated to English**. Relevant articles from the reference sections of included studies were also identified and added to the analysis. Historically, autistic individuals have been diagnosed with autistic disorder (AD) or Asperger’s syndrome (AS) depending on the severity of the autistic phenotype (Colleen & Stone, 2014). Present classification criteria make this subgrouping redundant, placing both groups under autism spectrum disorder (ASD). As several MRS autism studies predate this change in diagnostic approach, we included studies reporting cohorts of ASD, AD and AS diagnosis in the meta-analysis which explores autism. The impact of the explicit diagnosis will however be explored (ASD/AD/AS).

The initial search returned 3191 studies (29/09/22). 2345 studies were eligible for abstract screening after removal of duplicates. 2002 studies were excluded in the abstract screening (with full text screening performed if necessary). Six relevant articles from the reference sections of included studies were identified and manually added to the analysis. After initial abstract screening, 135 studies were eligible for full-text screening, with 76 excluded for the following reasons: no non-autistic control comparison (n = 12); not peer reviewed (n = 8), duplicated data (n = 10), not relevant (no MRS, MRSI used; n = 28), drug clinical trials with no recorded baseline (n = 4), the authors did not reply to requests for raw data and there were no other means of extracting raw data (n = 14). This is summarised in the PRIMSA chart (Figure 1).

**Figure 1.**
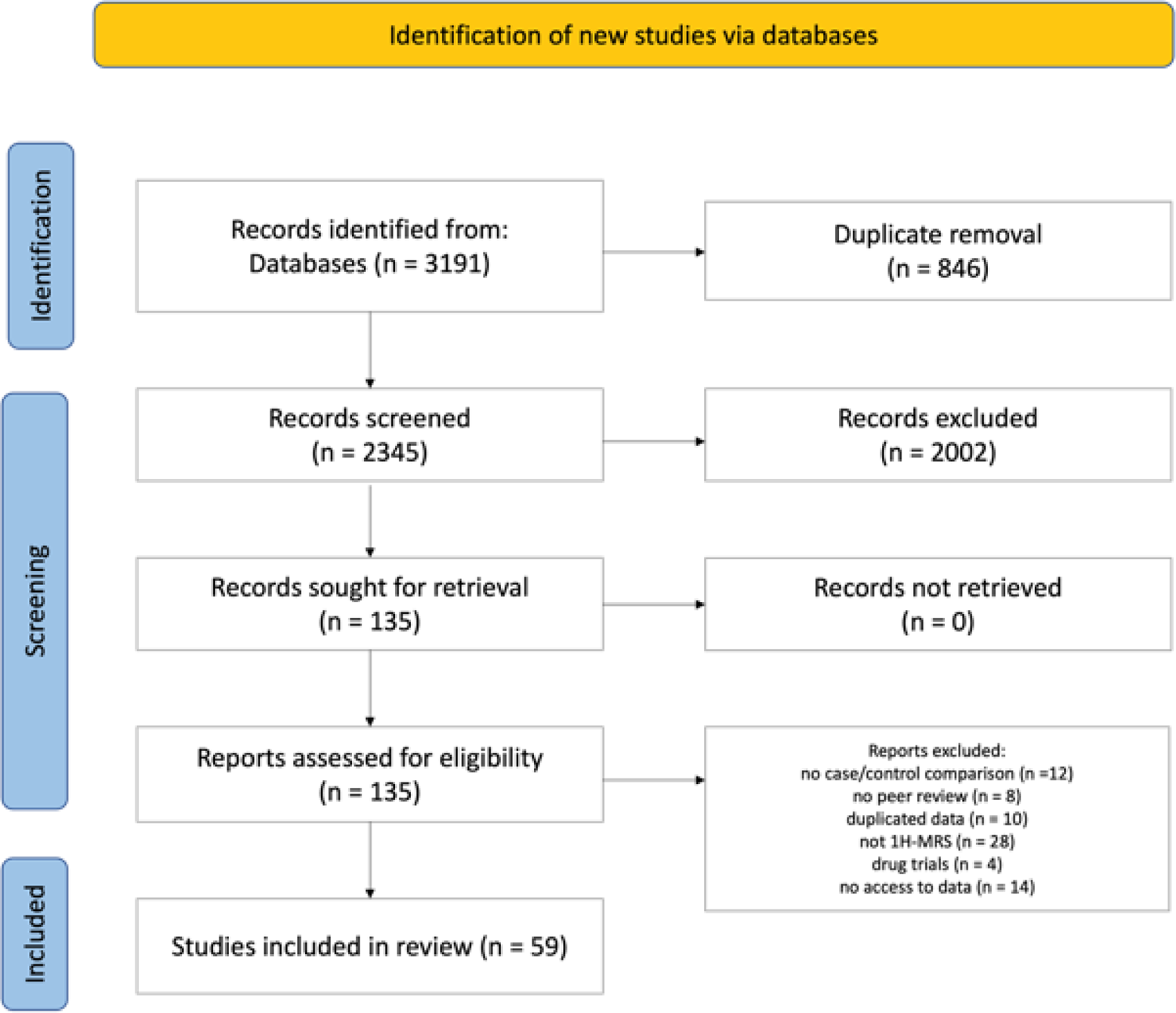
PRIMSA flow diagram of study selection. Papers were included if they met all of the following criteria: (1) use of in-vivo MRS to measure metabolites in the brain; (2) investigation of brain metabolite concentrations in autism and non-autism groups (3) MRS measured from a specified brain region and (4) the study was published in a peer-reviewed journal and was written in English or translated to English. Reasons for exclusion; duplicate removal (n = 846), records excluded as not relevant after abstract screening (n = 2002), no case/control comparison (n = 12), no peer review (n = 8), duplicated data (n = 10), not using MRS (n = 28), drug trails with no reported baseline (n = 4), no access to data after emailing (n = 14).

### 2.2 Study selection and data extraction table

Relevant abstracts were independently assessed by two investigators (NAP and AT). This was followed by full text screening, with reasons for exclusion documented. Data were then extracted in 4 domains 1) brain metabolite levels in non-autism and autism cohorts. We note that although we chose non-autistic and not neurotypical (NT) to define the control group, as some control participants may simply be non-autistic and less care was taken to exclude other neurodevelopmental conditions 2) study cohort characteristics (i.e., sample size, age, gender, IQ); 3) reported MRS acquisition parameters (e.g., MRS sequence, MRI scanner static field strength, echo time (TE)/ repetition time (TR), voxel size, averages, processing software, pre- and post-processing parameters); 4) bibliometric data (e.g., authors, year of publication, and type of publication). Key correlative findings were also noted for each study for systematic review. Data extraction table can be found in supplementary table 1, final data table used for analysis is available at https://osf.io/hxw3c/?view_only=5d2d683b027541879884485f138f5ddb.

Concentrations of GABA, Glx, Glu, Cr, tCho, NAA and mI were recorded from reported mean and standard deviation (SD) values in autism and non-autism groups. If raw data values were not reported, lead authors were emailed, and where this was not successful, imputation methods were used in accordance with recommendations in the Cochrane meta-analysis handbook (Higgins & Green, 2002). As an example of the latter, if the data was displayed in a figure only, WebPlotDigitizer (Rohatgi, 2022) was used to extract data manually.

Studies vary in their metabolite quantification method: articles can report concentrations of neurometabolites as a ratio of a stable reference metabolite measured within the same voxel, this usually being creatine. Alternatively, estimated neurometabolite concentrations in institutional units (i.u) are reported, whereby metabolite concentrations are calculated in reference to voxel water signal. As brain tissue types differ significantly in water composition, researchers should optimally control for differences in tissue composition between voxels and between groups by applying tissue correction. At the most basic level, metabolite concentrations are scaled according to the assumption that metabolite concentrations in CSF are negligible (Gasparovic et al. 2006; Harris et al., 2015). Additional corrections include ‘alpha correction’ of GABA concentrations, based on the assumption that GABA concentration is two times greater in grey matter (GM) compared to white matter (WM), as well as accounting for differences in voxel T1/T2 tissue relaxation (Jensen et al., 2005, Harris et al., 2015). We record if such corrections were performed for the relevant studies. Where studies reported estimated metabolite concentrations in i.u (water referenced) and creatine ratios, both were recorded, however as overall a greater number of studies reported concentrations in i.u, for the main analysis i.u was used in preference.

Across studies, specific metabolites were reported in various ways depending on the spectral separation achieved at each field strength, post-hoc processing corrections and assumptions made by individual researchers. For example, creatine was mostly reported as total creatine (tCr), this being the sum of creatine and its phosphorylated form phosphocreatine, as their peaks are hard to separate at 1.5/3 Tesla field strengths (Cr + pCr; N = 23). 4 articles reported creatine concentrations alone (Cr; N = 4). NAA was reported as total NAA; the sum of the overlapping peaks N-acetyl-aspartate (NAA) and N-acetyl-aspartyl-glutamate (NAAG; N = 3), or NAA alone (N = 32). Note if it was not stated if NAA signal was composed of both NAA and NAAG, it was assumed researchers reported NAA alone (N=15).

Finally, GABA was reported as GABA+ (GABA plus macromolecule contribution; N = 18), or as estimates of GABA concentrations alone (through using macromolecule suppression techniques/macromolecule basis set modelling; N = 8). Choline was always reported as a sum of the choline containing compounds; compounds phosphocholine, phosphatidylcholine and glycerophosphocholine (tCho; N = 27). Reports of GABA and GABA+ were grouped under “GABA”, reports of tNAA and NAA were grouped under “NAA” and reports of Cr + pCr and tCr were grouped under “creatine” for ease of analysis and to optimise inclusion for statistical power. Glx and Glu data was analysed separately.

### 2.3 Quality assessment

#### AXIS and MRS-Q

A modified appraisal tool for cross-sectional studies, specifically developed for MRS data, was used to assess cross sectional study design and reporting using 19 domains (AXIS; Downes et al., 2016; Peek et al., 2020). For each study, the percentage of AXIS domains satisfied was assessed. Domain 9 “were the risk factors and outcome variables measured correctly using instruments/measurements that had been trialled, piloted or published previously” of AXIS was discounted. Instead, the consensus quality appraisal tool ‘MRS-Q’ was used to comprehensively assess if MRS acquisition parameters satisfied consensus standards, and specifically, if reporting of MRS parameters was sufficient (A. Lin et al., 2021; Peek et al., 2020).

Studies that reported sufficient spectroscopy parameters to satisfy MRS-Q consensus standards for data acquisition were categorised as ‘high quality’ data. Studies that reported insufficient spectroscopy parameters to satisfy the consensus standards were classified as ‘low quality’ data, and studies with not enough information to make judgement were considered ‘unsure’. Importantly, this classification may fail to reflect the true quality of the study itself but reflects that reporting was poor. We do however analyse data with and without ‘low-quality’ studies to assess the impact of reported data quality on outcome. Two investigators (NAP and AT) independently assessed the quality of each study for both criterions.

### 2.4 Publication bias

Effect sizes were aggregated for each metabolite within each study (across the brain regions studied) using the ‘aggregate’ function from the metafor package in R (Viechtbauer, 2010). This attempted to increase the homogeneity of effect sizes and as such usefulness of publication bias tests, as well as preventing non-independence effects (due to multiple brain regions reported from the same participants in each study).

Egger’s regression tests (Bowden et al., 2015; Egger et al., 1997; Pasanta et al., 2023) were performed on aggregated metabolite data using the *meta* package function ‘metabias’ (Balduzzi et al., 2019). For Egger’s regression test (Egger et al., 1997; Sterne & Egger, 2001) a funnel plot was generated for each metabolite with effect size per study (across all brain regions) on the x axis and the standard error of the effect size on the y axis. In theory, without publication bias, effect sizes should follow a symmetrical distribution around 0 (L. Lin & Chu, 2018). Egger’s regression quantifies if effect sizes are asymmetrical and so may have a publication bias.

The trim-and-fill method is a non-parametric test used to correct for asymmetry of effect sizes due to publication bias (Duval & Tweedie, 2000; Pasanta et al., 2023). First, the studies driving asymmetry (extreme effect sizes) are identified and ‘trimmed’, and then a new estimate for the true centre (true effect size) of the distribution is found. This distribution is then ‘filled’ as the trimmed studies are added back, along with a prediction of the studies not reported due to publication bias (Balduzzi et al., 2019; Duval & Tweedie, 2000). For the trim-and-fill, a random-effects model using the Knapp and Hartung method was used on aggregated data using the *metabias* package *‘funnel.meta’* and *‘trimfill’* function (Balduzzi et al., 2019; Nakagawa et al., 2022; Pasanta et al., 2023). Data distribution was visualized on funnel plots (Begg & Mazumdar, 1994; Sterne & Egger, 2001).

### 2.5 Data analysis

Data-analysis was performed on extracted data using the *Meta-Essentials* tool in R (Suurmond et al., 2017). Standardized mean differences (Hedge’s G) and 95% confidence intervals were calculated from mean metabolite concentrations and standard deviation in the autism and non-autistic group (per study per metabolite), allowing us to compare data reported in different units (creatine scaled and i.u). Subgroup and meta-regression analyses (*robumeta* package) were used to explore if effect sizes were associated with demographic and methodological parameters. Note for all metabolites, mean non-autistic concentration was subtracted from mean autistic concentration and as such standardised mean differences (effect size) are expressed with respect to autism. For example, a negative effect size indicates lower concentrations of said metabolite in autism while a positive effect size indicates higher concentrations in autism compared to non-autistic controls.

Most studies acquired multiple voxels (brain regions) and metabolites from the same cohort, and so several effect size estimates were obtained from a single study e.g. NAA, Creatine and Glx from a frontal and temporal voxel per participant. This can lead to statistical dependency between measures, potentially skewing the calculation of group effect size parameters (Borenstein et al., 2021). To control for this, we use robust variance estimation (RVE) for group effect size calculations (Hedge’s g), using the *robumeta* package in R (Fisher & Tipton, 2015; Pasanta et al., 2023; Pustejovsky & Tipton, 2022). RVE assumes that effect sizes from the same study are correlated, and since the exact correlation is unknown, RVE estimates this dependence structure. A conservative correlation coefficient of 0.7 was used for all RVE calculations in accordance with (Pasanta et al., 2023; Rosenthal, 1986). Further adjustments of RVE estimates were made to control for small samples sizes (Tipton, 2015). Furthermore, when degrees of freedom where smaller than 4, a significance threshold of p < 0.01 was used (instead of p < 0.05) to assess significance of Hedge’s g, as type 1 errors are more likely in this case (Tanner-Smith et al., 2016).

Data heterogeneity was evaluated using I^2^ (Higgins et al., 2002) and Tau^2^ (Viechtbauer, 2005, 2010). I^2^ statistic is an estimate of the proportion of variance in effect sizes (between studies) that reflects true heterogeneity, ranging from 0 to 100. High I^2^ (>70), suggests there may be real differences between study outcomes not as a result of sampling error (instead due to real differences in study methods/cohort characteristics) and so sub-group analysis maybe more appropriate (Borenstein et al., 2021; Hak et al., 2016; Higgins & Green, 2002). Tau^2^ squared is the estimated variance of true effect sizes, in other words measuring the amount of true heterogeneity. If the confidence interval for tau^2^ does not contain 0, this suggests that between study heterogeneity exists and again subgrouping may be appropriate (Viechtbauer, 2005, 2010).

### 2.6 Subgrouping

Our main aim was to identify general differences in GABA, Glu, Glx, creatine, choline, GSH, NAA and mI concentrations between autistic and non-autistic cohorts and explore if this varies with age, brain region and study methodology. As such, we sub-grouped the data based on metabolite of interest, age group of cohorts (adults ≥ 18 years of age, children < 18 years of age) and brain region. We then grouped data by demographic factors (sex, IQ) and acquisition parameters (scanner strength, sedation, pulse sequence etc.,). We additionally looked at sex ratios in each study, testing both a 1:3 female:male ratio (as diagnostic bias in autism is currently 1:3 (Loomes et al., 2017), and 1:1 ratio for sex-balanced studies). The forestplot function in the *forestplot* package in R was used to plot RVE estimates of Hedges’ g per subgroup. We also used the *cor* function in r to estimate the Spearman’s rho correlation coefficient between voxel size/number of transients and study effect size.

To optimise study numbers per brain region, we grouped data into broader regions (brain region grouping 1; supplementary table 2) using previously defined and functionally relevant brain regions groupings (Pasanta et al., 2023). We use a secondary brain region grouping (brain region grouping 2; supplementary table 3) to explore significant findings with more spatial resolution, however study number per group was considerably reduced (e.g. n = 2-4) and as such brain grouping 1 was considered more appropriate and preferred for main interpretations.

Finally, as we aimed to explore whether there was an association between study outcome and quality of MRS data, we repeated the main analysis including only high-quality studies as defined by MRS-Q.

## 3. Results

### 3.1 MRS-Q

Most studies (n = 46/59, 88%) reported methods that satisfied the MRS-Q criteria of best practice and so were deemed to be of high quality (Figure 2). Ten studies (17%) were assessed as low quality; this was mostly due to small voxel sizes and too few transients. All low-quality studies used conventional (non-editing) sequences (so were not observing GABA; Bernardi et al., 2011; Goji et al., 2017; Harada et al., 2011; Hardan et al., 2008, 2016; Otsuka et al., 1999), while 2 also used edited MRS sequences in combination with conventional MRS (Goji et al., 2017; Harada et al., 2011). Note for (Goji et al., 2017) and (Harada et al., 2011) only conventional (non-editing) MRS data was classified as low quality, edited MRS acquisition parameters were assessed separately and classified as high quality. For the high quality studies, 27 studies used conventional (non-editing) MRS sequences (L. Lin & Chu, 2018). 12 used editing MRS sequences (Bernardino et al., 2022; DeMayo et al., 2021; Drenthen et al., 2016; Fung et al., 2021; Gaetz et al., 2014; Kirkovski et al., 2018; Pierce et al., 2021; Port et al., 2019; Puts et al., 2017; Rojas et al., 2014; Sapey-Triomphe et al., 2021; Umesawa et al., 2020). 8 studies used both editing and conventional (non-editing) sequences (Brix et al., 2015; Carvalho Pereira et al., 2018; Cochran et al., 2015; He et al., 2021; Ito et al., 2017; Maier et al., 2022; Moxon-Emre et al., 2021; Pierce et al., 2021; Wood-Downie et al., 2021). Three studies were classed as unsure due to missing information regarding MRS parameters (Drenthen et al., 2016; Kubas et al., 2012; Tebartz van Elst et al., 2014).

**Figure 2:**
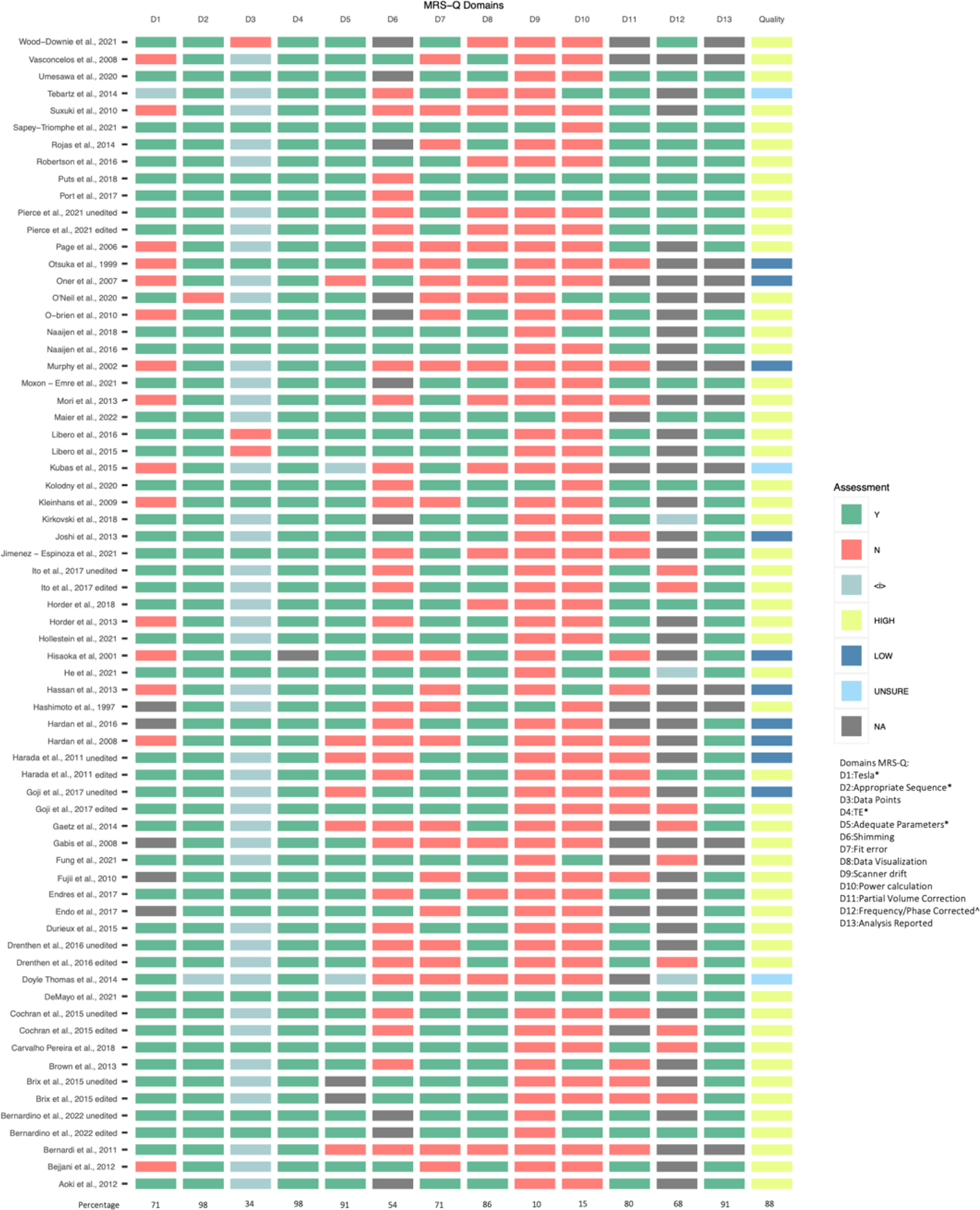
MRS quality as assessed by MRS-Q. MRS-Q assessment of MRS studies, domain titles are shown in the key. * Indicates the domain was used for quality assessment of MRS acquisition and reporting. ^ indicates that this domain is only applicable to edited MRS.

The poorest performing domains for MRS-Q were: reporting of scanner drift (only 10% of studies), reporting of number of MRS data points (30% of studies) and reporting of power calculations for sample size/justification of chosen sample size (only 16% of studies).

### 3.2 AXIS

While on average the included studies satisfied 68% of the AXIS criteria (Figure 3), the quality of the studies varied. Eight studies satisfied over 80% of the criteria (Bernardino et al., 2022; Brix et al., 2015; DeMayo et al., 2021; Durieux et al., 2016; Gaetz et al., 2014; Hollestein et al., 2021; Horder et al., 2018; Suzuki et al., 2010), while 10 studies satisfied 50% or less of the criteria only (Gabis et al., 2008; Harada et al., 2011; Hashimoto et al., 1997; Hisaoka et al., 2001; Kirkovski et al., 2018; Kubas et al., 2012; Oner et al., 2007; Otsuka et al., 1999; Page et al., 2006; Robertson et al., 2016). 96% of studies clearly defined an aim, the ‘case’ population and included sufficient methods for replication, however, only 26% of studies selected a representative control population from the same local population as case populations, only 56% of studies categorized non responders and 52% justified excluded scans. Studies also varied in controlling for confounding factors; 54% explicitly stated that co-occurring conditions were excluded, while only 24% of studies excluded or adjusted for medication use. Other confounders such as sedation, IQ, and age were mostly controlled, mostly post-hoc statically (80% of studies). As we extract raw data, these post-hoc statistical controls do not stand in our meta-analysis. Most studies (around 70%) presented all results and justified their conclusions.

**Figure 3:**
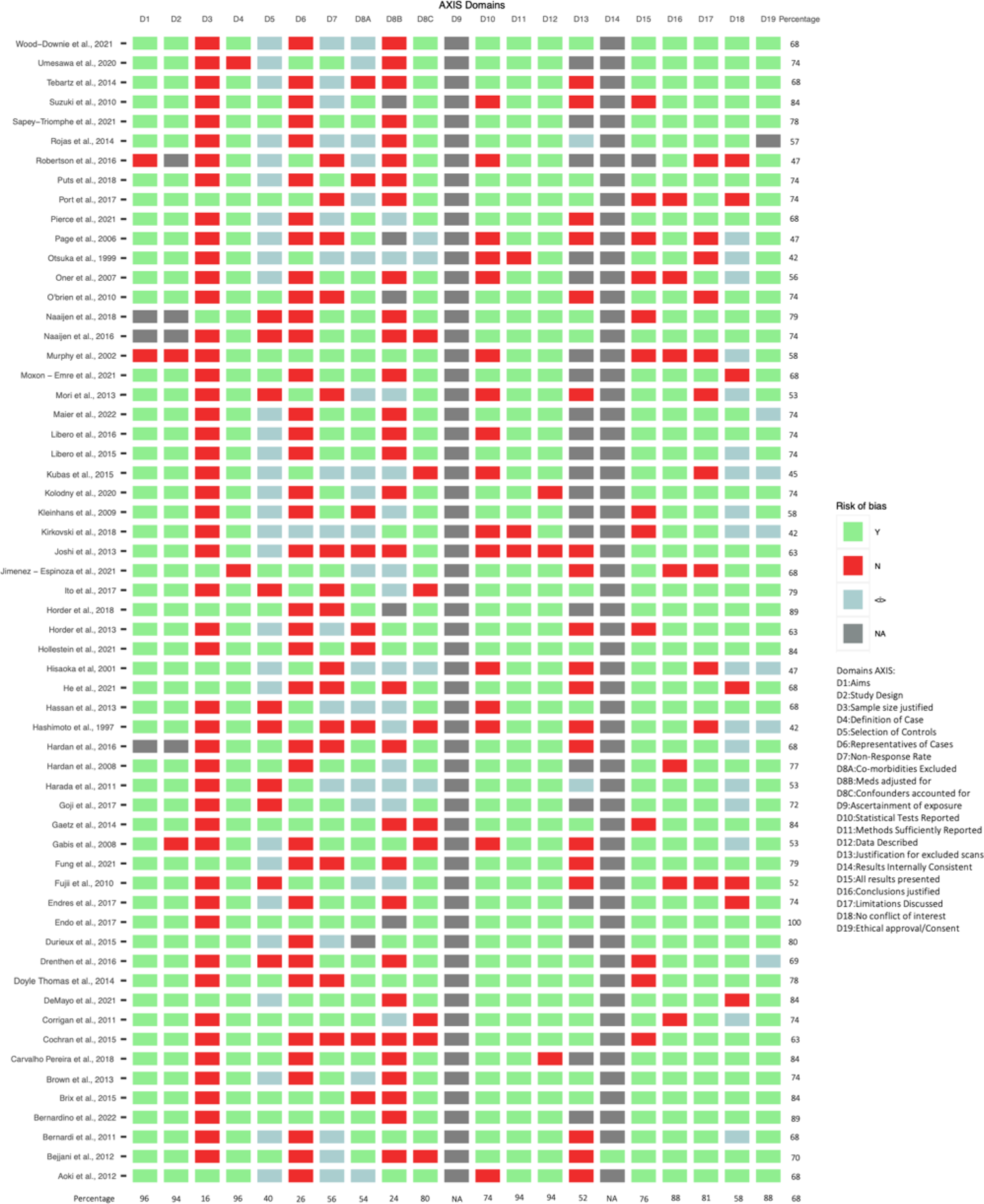
Study design assessed by AXIS. A modified appraisal tool for cross-sectional studies (AXIS), specifically developed for MRS, was used to assess the quality of MRS study design and reporting.

### 3.3 Publication bias assessment

#### Egger’s regression test

No significant funnel plot asymmetry was found for NAA, choline, creatine, Glx, Glu and mI (Table 1). Significant asymmetry (p < 0.05) was identified for studies observing GABA. Funnel plots suggest an absence of positive effect sizes. Note all GABA studies with a large negative effect size, and so driving this asymmetry, reported creatine-scaled metabolite concentrations (Gaetz et al., 2014; Kubas et al., 2012; Port et al., 2019). For GSH, too few studies had been performed (n = 3) for appropriate bias assessment.

### Trim-and-fill

Three studies were ‘trimmed and filled’ for NAA (Gabis et al., 2008; Hardan et al., 2016; Kubas et al., 2012) and 2 for mI (Brown et al., 2013; Gabis et al., 2008), with the resulting estimated effect sizes reported in Table 3.

**Table 2.** Egger’s Regression test for publication bias (funnel plot asymmetry).

| Metabolite | Bias | Se | Intercept | Se | t | p | df | Result |
| --- | --- | --- | --- | --- | --- | --- | --- | --- |
| NAA | -2.0934 | 1.1143 | 0.3898 | 0.3474 | -1.88 | 0.0692 | 33 | No bias |
| choline | 0.8477 | 0.8527 | -0.2251 | 0.2625 | 0.99 | 0.3297 | 25 | No bias |
| creatine | -1.4758 | 1.5770 | 0.3576 | 0.5106 | -0.94 | 0.3583 | 25 | No Bias |
| Glx | -0.9139 | 1.3760 | 0.2388 | 0.4202 | -0.64 | 0.5301 | 27 | No Bias |
| Glu | 0.4854 | 1.4268 | -0.0136 | 0.3854 | 0.34 | 0.7401 | 11 | No Bias |
| GABA | -3.1769 | 1.2545 | 0.6360 | 0.3785 | -2.53 | 0.0183 | 24 | Bias |
| GSH | NA | NA | NA | NA | NA | NA | NA | NA |
| ml | 0.4494 | 1.4071 | -0.2171 | 0.5464 | 0.32 | 0.7531 | 18 | No Bias |

Note: significance indicates an asymmetrical distribution of effect sizes on funnel plots.

**Table 3.** Trim-and-Fill correction for publication bias.

| Metabolite | Filled | Adjusted<br>Estimate<br>effect size | 95%CI | T value | p | tau<sup>2</sup> |
| --- | --- | --- | --- | --- | --- | --- |
| NAA | Kubas et al., 2012 |  |  |  |  |  |
|  | Gabis et al., 2008 | -0.1771 | -0.5230; 0.1689 | -1.04 | 0.3064 | 0.7710 |
|  | Hardan et al., 2016 |  |  |  |  |  |
| choline | None | 0.0310 | -0.1137; 0.1756 | 0.44 | 0.6636 | 0.0115 |
| creatine | None | -0.1369 | -0.4039; 0.1301 | -1.05 | 0.3015 | 0.2630 |
| Glx | None | -0.0417 | -0.3094; 0.2260 | -0.32 | 0.7519 | 0.2868 |
| Glu | None | 0.1086 | -0.2179; 0.4351 | 0.72 | 0.4825 | 0.1230 |
| GABA | None | -0.4124 | -0.7505; -0.0743 | -2.51 | 0.0188 | 0.4351 |
| GSH | None | 0.0962 | -1.3191; 1.5115 | 0.29 | 0.7975 | 0.2318 |
| MI | Brown et al., 2012 |  |  |  |  |  |
|  | Gabis et al., 2008 | -0.1914 | -0.6107; 0.2278 | -0.95 | 0.3531 | 0.5973 |

All data demonstrated high heterogeneity, with I^2^ calculated at 70-80% even when data was split by metabolite (Figure 3). This suggests that more than half of the variation in estimated effect sizes is due to true differences in effect size between studies, likely because of the differences in brain region, age etc. It has been advised in such cases publication bias analysis may not be appropriate (Nakagawa et al., 2022; van Aert et al., 2016). As such, identified ‘trimmed’ studies remain in the analysis, while publication bias results were considered during interpretation.

### 3.4 Study characteristics

All study characteristics are shown in Table 4.

**Table 4.**
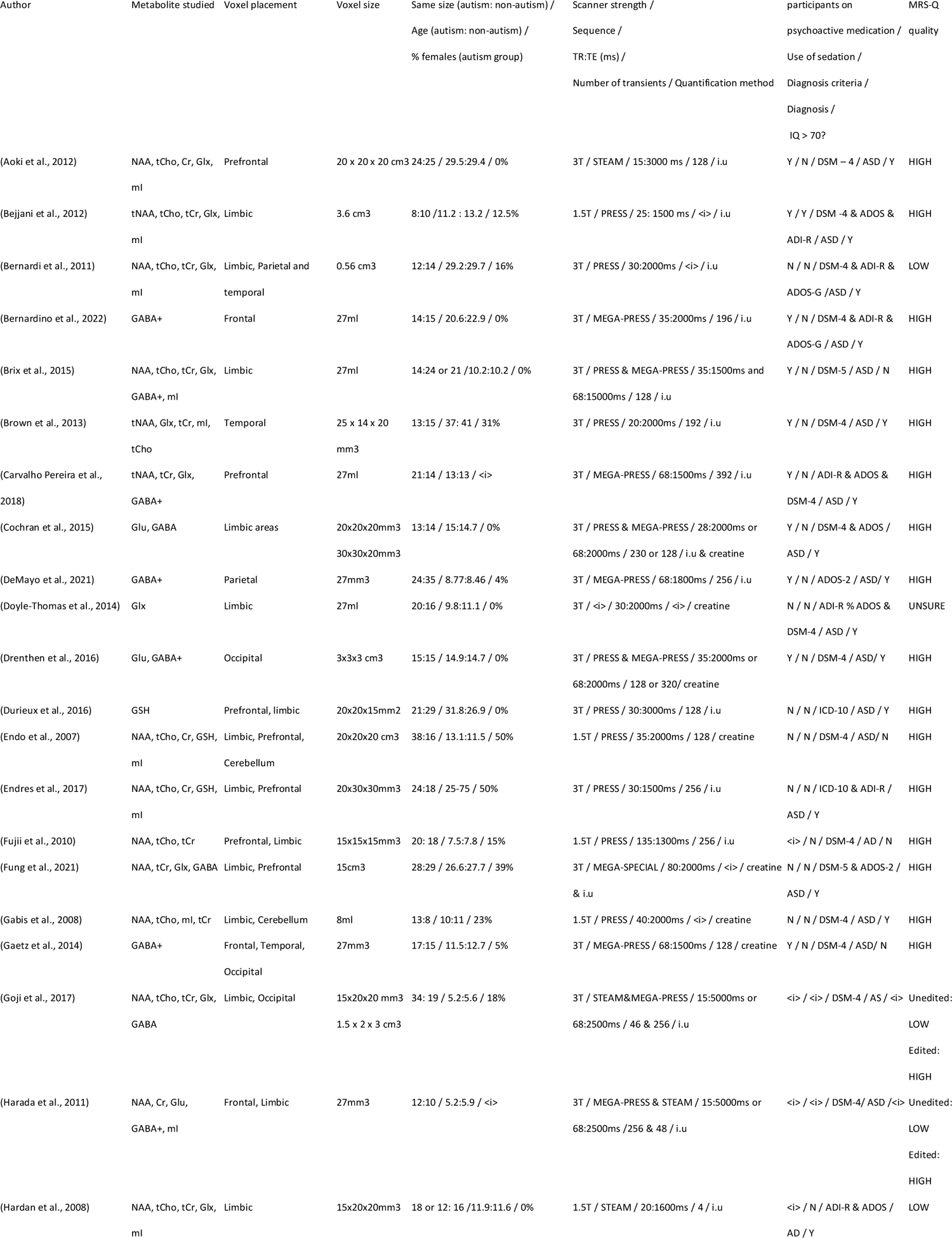
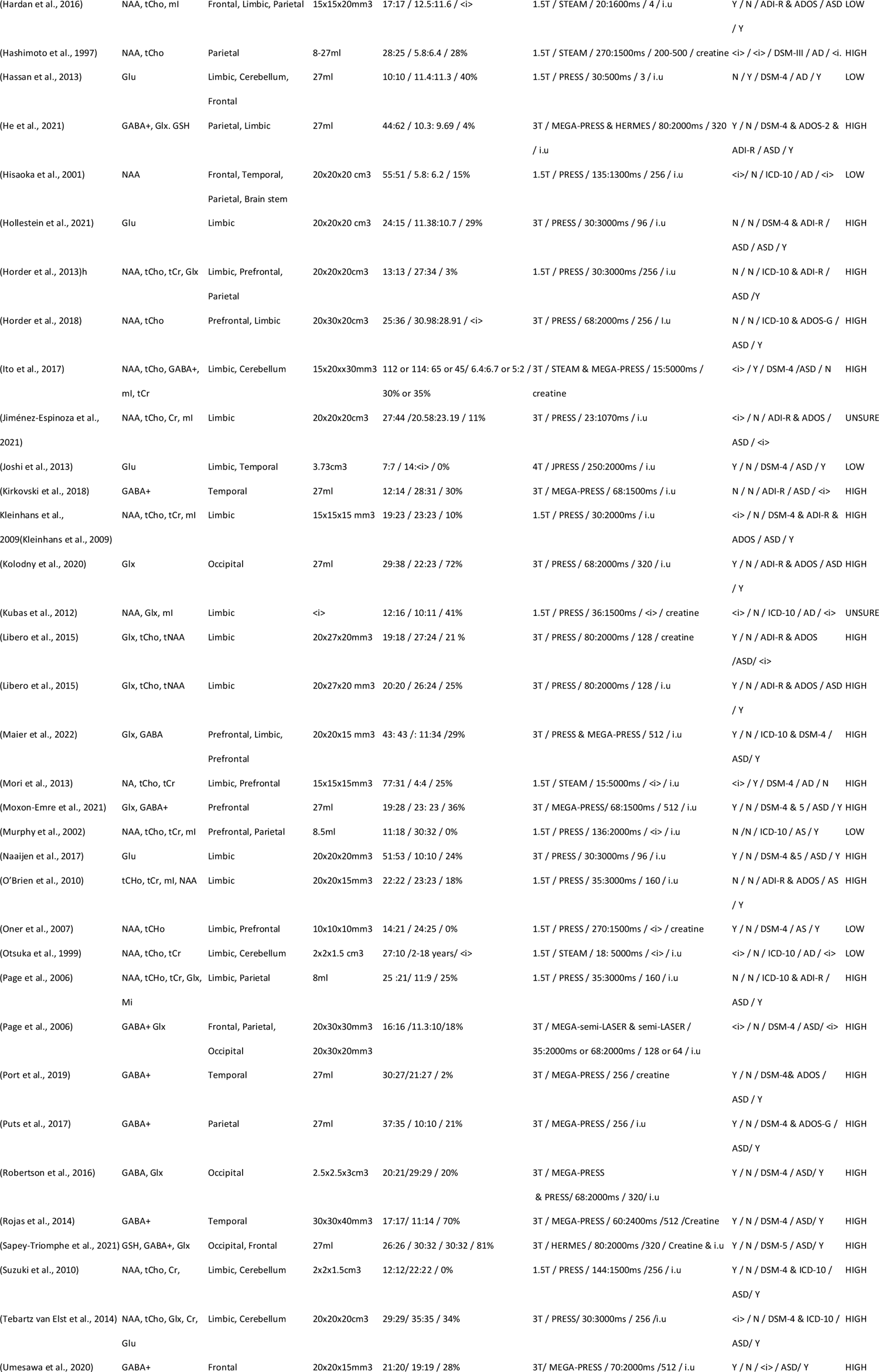
Study characteristics.

### 3.5 Meta-analysis results

Note all group effect sizes reported below are in respect to autism, as such a negative effect size suggests lower metabolite concentrations in autism and a positive effect size suggests higher metabolite concentrations in autism.

#### General trends

We first considered the change in concentration across all metabolites, brain regions, age groups and other factors (e.g., pulse sequence; Figure 4). We found a non-significant negative effect size (Hedges’ g = −0.16, 95% CI: −0.34 − 0.022, I^2^ = 82, n = 59, 57.6 % children, p = 0.085).

**Figure 4.**
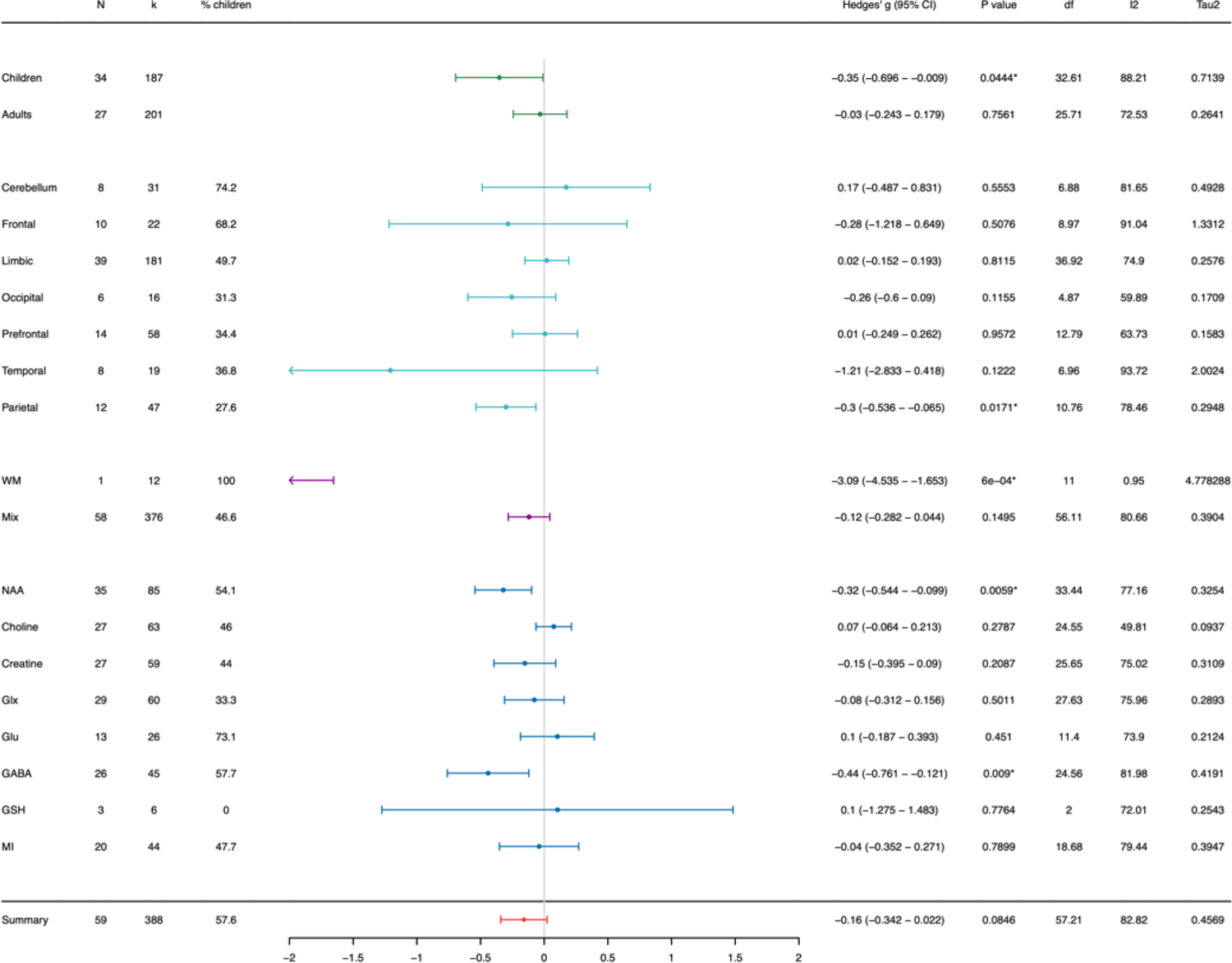
Summary forest plot for data grouped by age, brain region, voxel tissue type, and metabolite of interest. N: number of studies, k: number of observations, % children: percentage of studies observing metabolite concentrations in children only cohorts, I^2^: measure of between study heterogeneity, Tau^2^: Variance in true effect sizes (another measure of between study heterogeneity). Hedge’s g is reported in respect to autism. *Statistically significant at p < 0.05, and at p < 0.01 when the degrees of freedom < 4 for RVE t-tests.

#### Grouped by age group

Based on reported age range of cohorts, 25 studies reported adult only cohorts (≥ 18 years) and 32 studies reported child cohorts (< 18 years). Note three studies included cohorts with a minimum age of 16/17 years, although had population mean age greater than 18 years and so cohorts were classified as adult (Bernardino et al., 2022, mean age = 29 years, range: 17 – 30 years; Jiménez-Espinoza et al., 2021, mean age = 20 years, range: unknown; Moxon-Emre et al., 2021, mean age = 23 years, range: 16 – 34 year).

When data were grouped by cohort age (adults or children), across all metabolites and brain regions (Figure 4), we observed a significant negative group effect size for studies observing metabolites in **children** (Hedges’ g = −0.35, 95% CI: −0.70 − −0.009, I^2^ = 88, n = 34, p < 0.05). In **adults**, we observed a non-significant group effect size close to zero (Hedges’ g = −0.03, 95% CI: −0.24 − 0.18, I^2^ = 72, n = 27, p = 0.76).

#### Grouped by brain region

30 studies took measurements from more than one brain region. The most reported brain area (brain region grouping 1) was the limbic regions (39 studies; 181 observations), followed by the prefrontal regions (14 studies, 58 observations), the parietal regions (12 studies, 47 observations), the frontal regions (10 studies, 22 observations), the cerebellum (8 studies, 31 observations), temporal regions (8 studies, 19 observations) and occipital regions (6 studies, 16 observations).

When data were grouped by brain region (grouping 1, across all metabolites and ages; Figure 4). We found a significant (negative) group effect size for studies observing metabolites in the parietal regions only (Hedges’ g = −0.30, 95% CI: −0.54 − −0.065, I^2^ = 78, n = 12, p < 0.05).

#### Grouped by metabolite

43 studies observed more than one metabolite. NAA was the most studied metabolite (35 studies), followed by Glx (29 studies), choline (27 studies), creatine (27 studies), GABA (26 studies), mI (20 studies), Glu (13 studies) and GSH (3 studies). When the data were grouped by metabolite (across all brain regions and ages; Figure 4), group effect sizes for NAA and GABA were significant and negative (Hedges’ g_NAA_ = −0.32, 95% CI: −0.54 − −0.099, I^2^ = 77, n = 35, 54.1 % children, p < 0.05 and Hedges’ gGABA = −0.44, 95% CI: −0.76 − −0.12, I^2^ = 82, n =26, 57.8 % children, p < 0.05).

#### Grouped by voxel tissue type

Data were also grouped by tissue type (WM/ GM/mixed) as one study reported metabolite concentrations in WM only. The majority (58) did not explicitly state the tissue type, and as such it was assumed that metabolite concentrations were assessed across the whole voxel (WM and GM), although voxel tissue composition likely varied greatly depending on brain region observed. The one study observing metabolite concentrations in WM only had a significant negative effect size (Hedges’ g = −3.09, 95% CI: −4.54 − −1.65, I^2^ = 0.95, n = 1, 100% children, p < 0.01; Figure 4). Across all brain regions, metabolites and ages, the other 58 studies that used mixed voxels (WM and GM) had a non-significant negative effect size (Hedges’ g = −0.12, 95% CI: −0.28 – 0.28, I^2^ = 56, n = 58, 46.6% children, p = 0.15; Figure 4).

To observe the effect of metabolite, brain region and age group simultaneously on effect sizes, data were split by:

1. metabolite of interest and brain region (Figure 5).
2. metabolite of interest and age group (Figure 6).
3. metabolite of interest, age group, and brain region (Figure 7-13).

**Figure 5.**
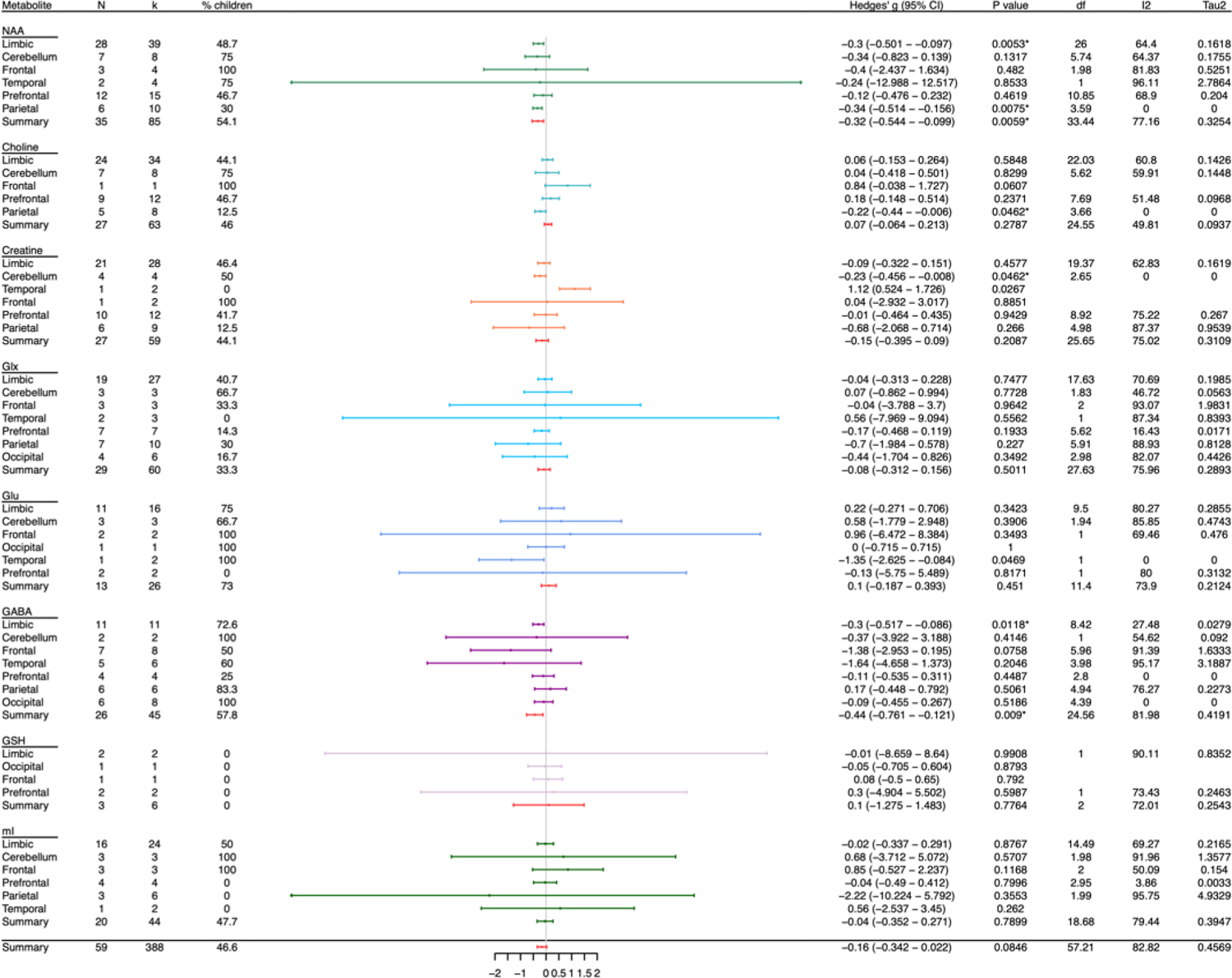
Summary forest plot for data grouped by metabolite and brain region. N: number of studies, k: number of observations, % children: percentage of studies observing children only, I^2^: measure of between study heterogeneity, Tau^2^: Variance in true effect sizes (another measure of between study heterogeneity). Hedge’s g is reported in respect to autism. *Statistically significant at p < 0.05, and at p < 0.01 when the degrees of freedom < 4 for RVE t-tests.

**Figure 6.**
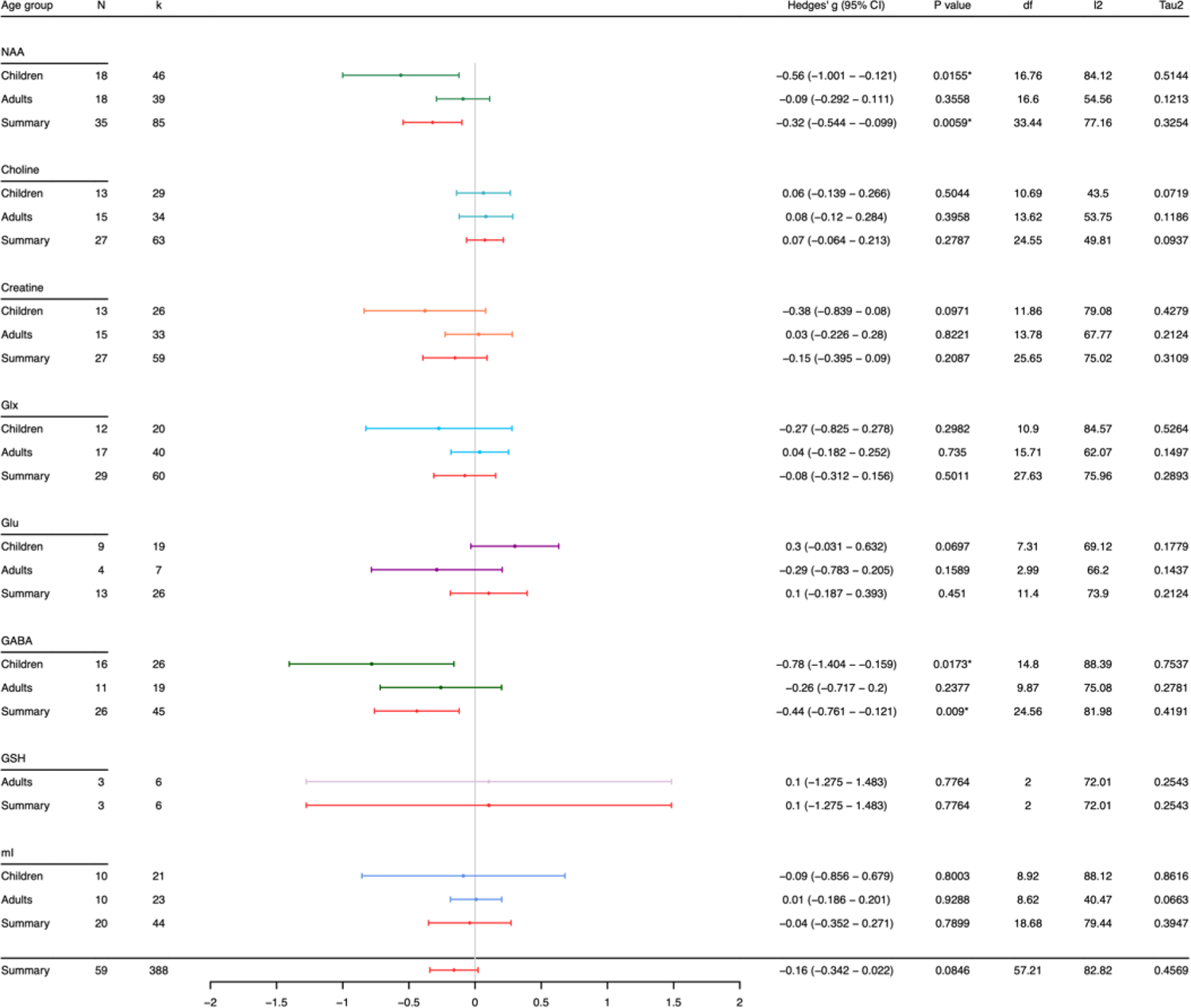
Summary forest plot for data by metabolite and age group. N: number of studies, k: number of observations, % children: percentage of studies observing children only, I^2^: measure of between study heterogeneity, Tau^2^: Variance in true effect sizes (another measure of between study heterogeneity). Hedge’s g is reported in respect to autism. *Statistically significant at p < 0.05, and at p < 0.01 when the degrees of freedom < 4 for RVE t-tests.

**Figure 7.**
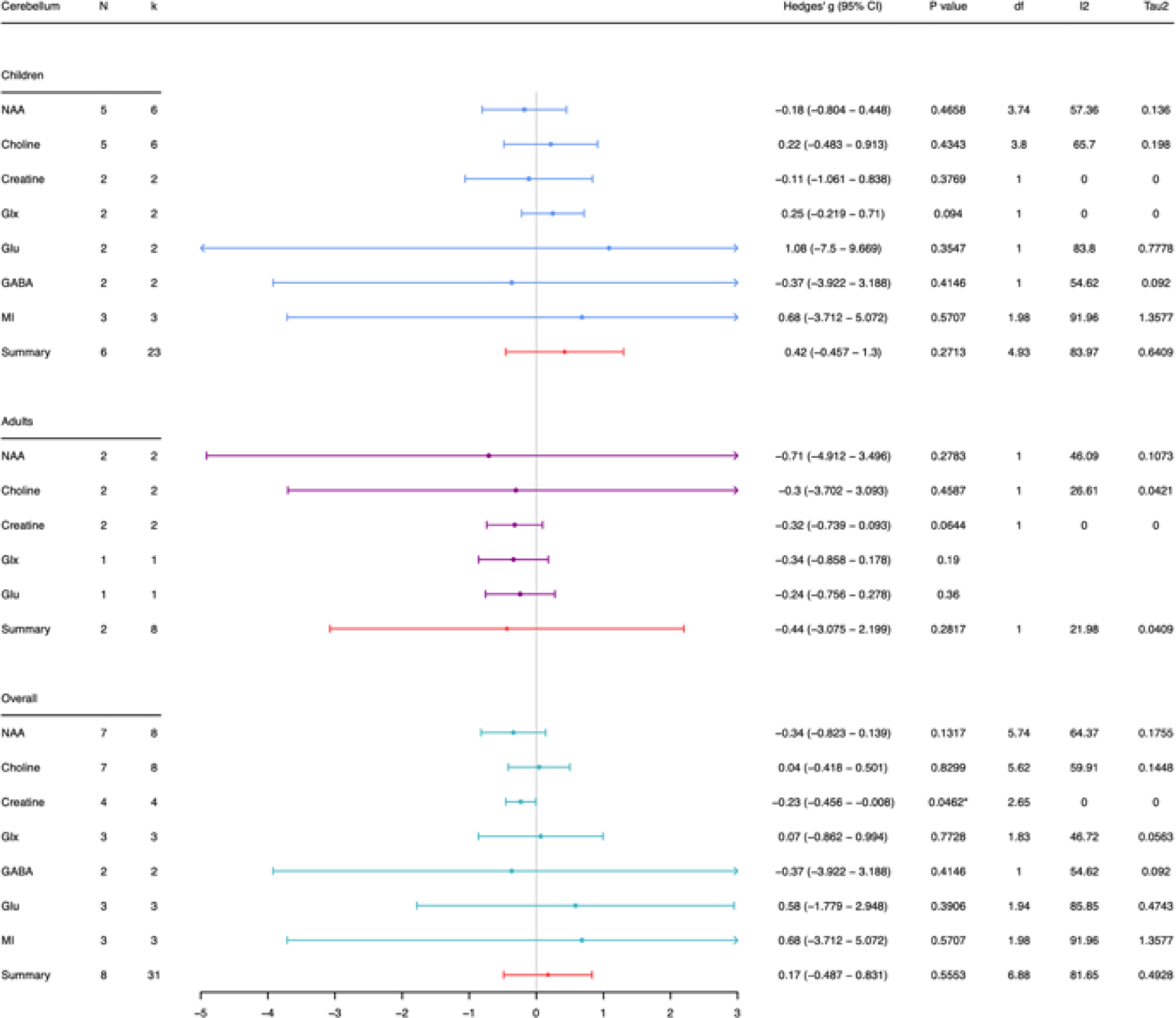
Summary forest plot findings in the cerebellum split by metabolite and age group (children, adults, and pooled ages (overall)). N: number of studies, k: number of observations, % children: percentage of studies observing children only, I^2^: measure of between study heterogeneity, Tau^2^: Variance in true effect sizes (another measure of between study heterogeneity). Hedge’s g is reported in respect to autism. *Statistically significant at p < 0.05, and at p < 0.01 when the degrees of freedom < 4 for RVE t-tests.

Results for these subgroupings are reported per metabolite below. Note if n = 1 for any subgroup explored, these results will be reported but excluded from interpretation.

#### NAA

*By brain region* A significant negative group effect size was found for studies measuring NAA in **limbic** (Hedges’ g = −0.3, 95% CI: −0.50 − −0.097, I^2^ = 64, n = 28, p < 0.05, 48.7% children; Figure 5) and **parietal** brain regions (Hedges’ g = −0.34, 95% CI: −0.51 − −0.16, I^2^ = 0, n = 6, p < 0.05, 30% children; Figure 5). No significant effect sizes were observed for studies measuring NAA in the cerebellum (n = 7; 75% children), frontal (n = 3; 100% children), temporal (n = 2; 75% children) or prefrontal regions (n = 12; 46.7 % children).

To explore this in more spatial detail we grouped NAA data by brain region grouping 2, where limbic regions are differentiated. We observed a significant negative effect size in the ACC only (supplementary figure 2; Hedges’ g = −0.22, 95% CI: −0.43 − −0.01, I^2^ = 41.5, n = 14, p < 0.05). Note this region had considerably greater number of studies than all other limbic brain regions (basal ganglia, PCC, medial temporal lobe (hypothalamus-amygdala)).

*By age group* A significant negative group effect size was observed for studies measuring NAA in **children** (Hedges’ g = −0.56, 95% CI: −1.00 − −0.12, I^2^ = 84, n = 18, p <0.05; Figure 6) but not in **adults** (Hedges’ g = −0.09, 95% CI: −0.29 − 0.11, I^2^ = 55, n = 18, p = 0.36; Figure 6).

*By age group and brain region* When studies measuring NAA where split by brain region and age group (Figure 7 – 13), a significant negative group effect size was observed for studies measuring **limbic** and **prefrontal** NAA in **children** (Hedges’ glimbic = −0.42, 95% CI: −0.80 − − 0.041, I^2^ = 75, n = 15, p <0.05; Figure 9 and Hedges’ g_prefrontal_ = −0.58, 95% CI: −1.10 − −0.067, I^2^ = 18, n = 4, p <0.05; Figure 10) but not in **adults** (Hedges’ g_limbic_ = −0.16, 95% CI: −0.37 − 0.065, I^2^ = 39, n = 14, p = 0.15; Figure 9 and Hedges’ g_prefrontal_ = 0.11, 95% CI: −0.3 − 0.51, I^2^ = 59, n = 8, p = 0.56; Figure 10).

For the cerebellum (n_children_ = 5, n_adult_ = 2; Figure 7), frontal (n_children_ =3, n_adult_ = 0; Figure 8), parietal (n_children_ =3, n_adult_ = 4; Figure 11), and temporal regions (n_children_ = 1, n_adult_ = 1; Figure 12) no significant effect size was observed for either age group for NAA.

**Figure 8.**
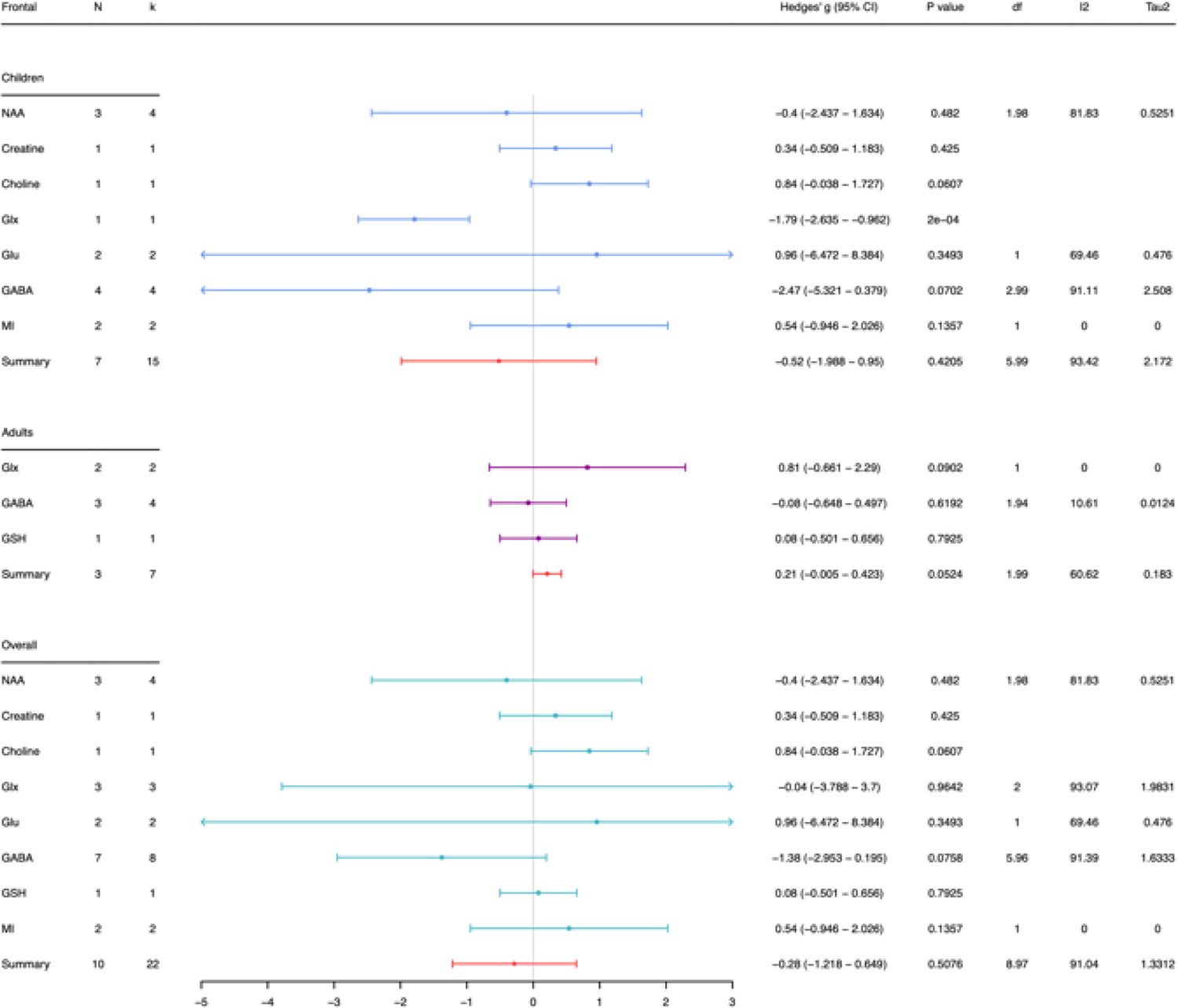
Summary forest plot findings in the frontal region split by metabolite and age group (children, adults, and pooled ages (overall)). N: number of studies, k: number of observations, % children: percentage of studies observing children only, I^2^: measure of between study heterogeneity, Tau^2^: Variance in true effect sizes (another measure of between study heterogeneity). Hedge’s g is reported in respect to autism. *Statistically significant at p < 0.05, and at p < 0.01 when the degrees of freedom < 4 for RVE t-tests.

To explore significant limbic findings in more detail, we grouped data by brain region grouping 2 (where limbic regions are differentiated) and age group. The ACC was the only limbic region with sufficient study numbers to explore age effects. We observed no significant effect size for ACC NAA concentrations in children (Hedges’ g = −0.11, 95% CI: − 0.40 – 0.17, I^2^ = 33, n = 7, p = 0.36; supplementary figure 3) or adult (Hedges’ g = −0.34, 95% CI: −0.71 – 0.03, I^2^ = 41, n = 7, p = 0.65). ACC NAA findings are only significant when adult and child data is collated (Supplementary figure 2; Hedges’ g = −0.22, 95% CI: −0.43 − −0.01, I^2^ = 41.5, n = 14, p < 0.05).

### GABA

*By brain region* We observed a significant negative group effect size for studies measuring GABA in the **limbic** regions (Hedges’ g = −0.3, 95% CI: −0.53 − −0.086, I^2^ = 27, n = 11, p < 0.05, 72.7% children; Figure 5). No significant effect size was observed for GABA in the cerebellum (n = 2; 100% children), frontal (n = 7; 50% children), temporal (n = 60% children), prefrontal (n = 4; 25% children), parietal (n = 6; 83% children) and occipital regions (n = 6; 100% children).

To explore this in more detail we grouped GABA data using brain region grouping 2, where limbic regions are differentiated. We observed a negative effect size in the ACC only, however this did not withstand a more stringent p value threshold of p < 0.01 due to df < 4 (Supplementary figure 2; Hedges’ g = −0.4, 95% CI: −0.74 − −0.065, I^2^ = 0, n = 5, p < 0.05, df = 2.6). Note for other limbic regions (thalamus, striatum) study number was small (n=2-3) and effect sizes were not significant.

*By age group* A significant negative group effect size was observed for studies measuring GABA in **children** (Hedges’ g = −0.78, 95% CI: −1.40 − −0.16, I^2^ = 88, n = 16, p <0.05; Figure 6) but not **adults** (Hedges’ g = −0.26, 95% CI: −0.72 − 0.2, I^2^ = 75, n = 11, p = 0.24; Figure 6).

*By age group and brain region* When studies measuring GABA where split by brain region (grouping 1) and age group (Figure 7 – 13), a significant negative group effect size was observed for **limbic** GABA in **children** (Hedges’ g = −0.35, 95% CI: −0.69 − −0.011, I^2^ = 45, n = 8, p < 0.05; Figure 9). This was also observed in **adults** when p < 0.05, however due to df < 4, a conservative p < 0.01 was used and significance was lost (Hedges’ g = −0.2, 95% CI: −0.36 − − 0.045, I^2^ = 0, n = 3, p = 0.0324 (df < 4); Figure 9).

**Figure 9.**
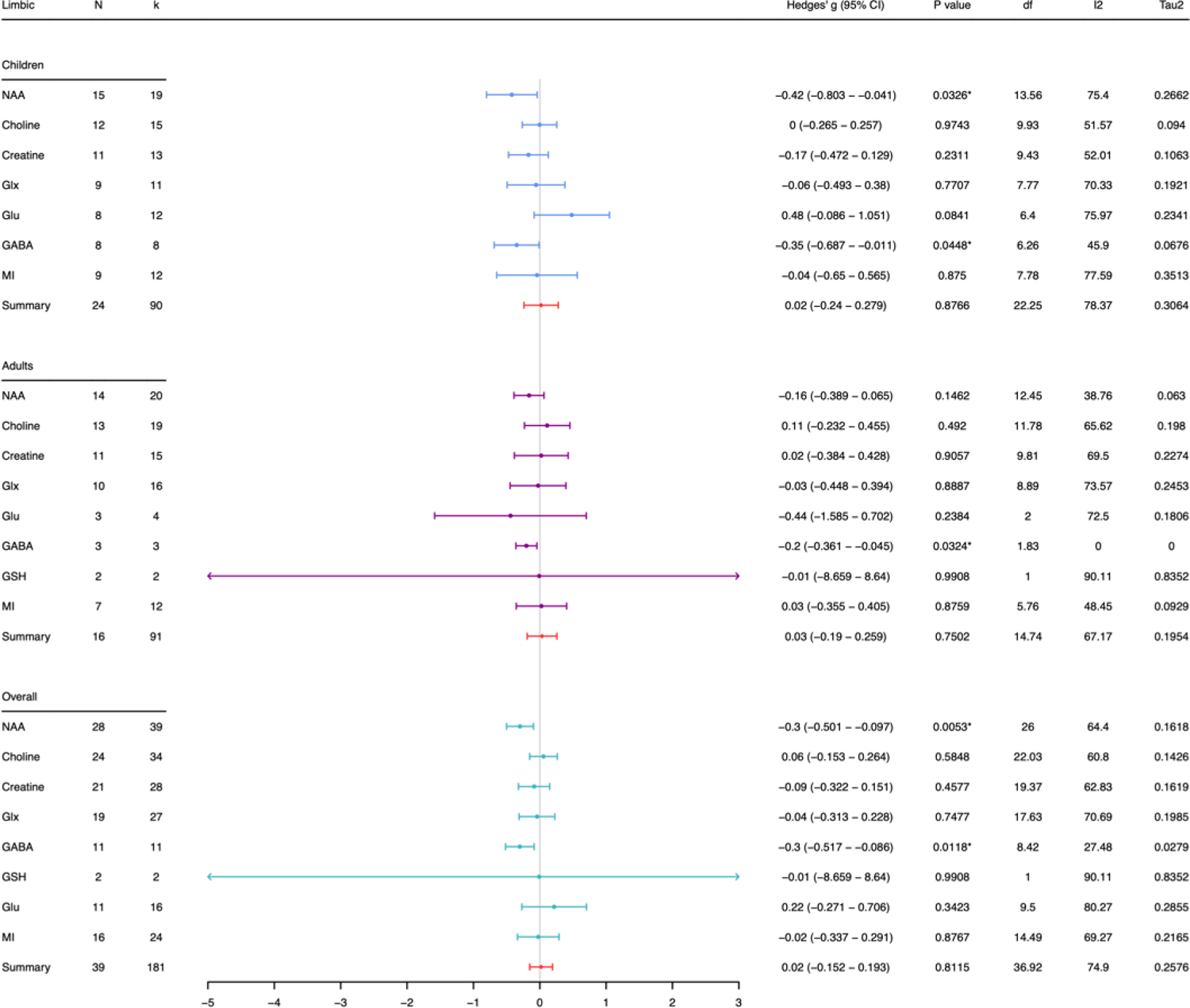
Summary forest plot findings in the limbic regions split by metabolite and age group (children, adults, and pooled ages (overall)). N: number of studies, k: number of observations, % children: percentage of studies observing children only, I^2^: measure of between study heterogeneity, Tau^2^: Variance in true effect sizes (another measure of between study heterogeneity). Hedge’s g is reported in respect to autism. *Statistically significant at p < 0.05, and at p < 0.01 when the degrees of freedom < 4 for RVE t-tests.

**Figure 10.**
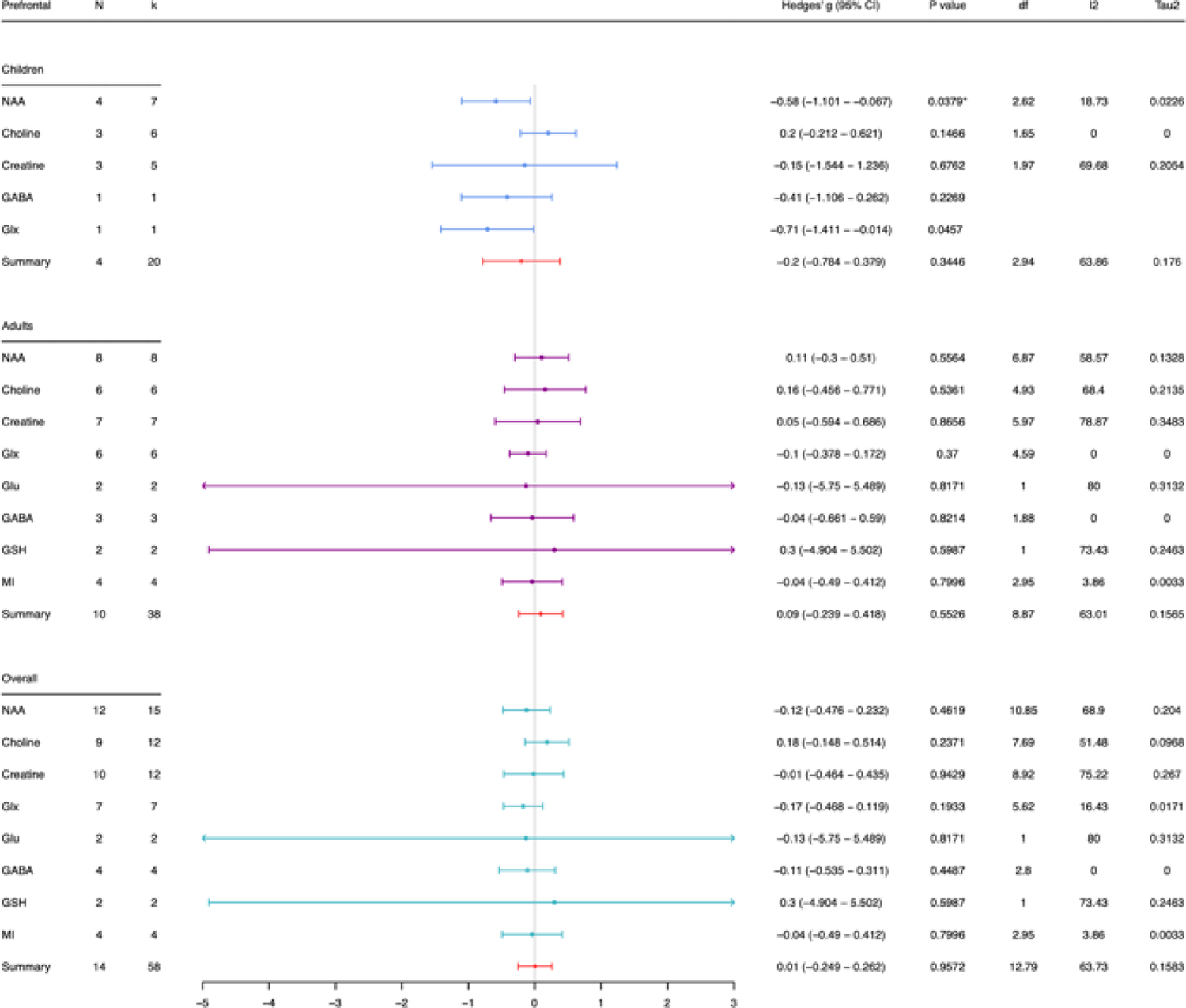
Summary forest plot findings in the prefrontal region split by metabolite and age group (children, adults, and pooled ages (overall)). N: number of studies, I^2^: measure of between study heterogeneity, Tau^2^: Variance in true effect sizes (another measure of between study heterogeneity). Hedge’s g is reported in respect to autism. *Statistically significant at p < 0.05, and at p < 0.01 when the degrees of freedom < 4 for RVE t-tests.

For studies measuring GABA in the cerebellum (nchildren = 2, nadult = 0; Figure 7), frontal (n_children_ = 4, n_adult_ = 3; Figure 8), prefrontal (n_children_ =1, n_adult_ = 3; Figure 10), parietal (n_children_ =5, n_adult_ = 1; Figure 11), temporal (n_children_ =3, n_adult_ = 0; Figure 12) and occipital regions (n_children_ = 3, n_adult_ = 3; Figure 13) no significant effect size was observed for either age group.

**Figure 11.**
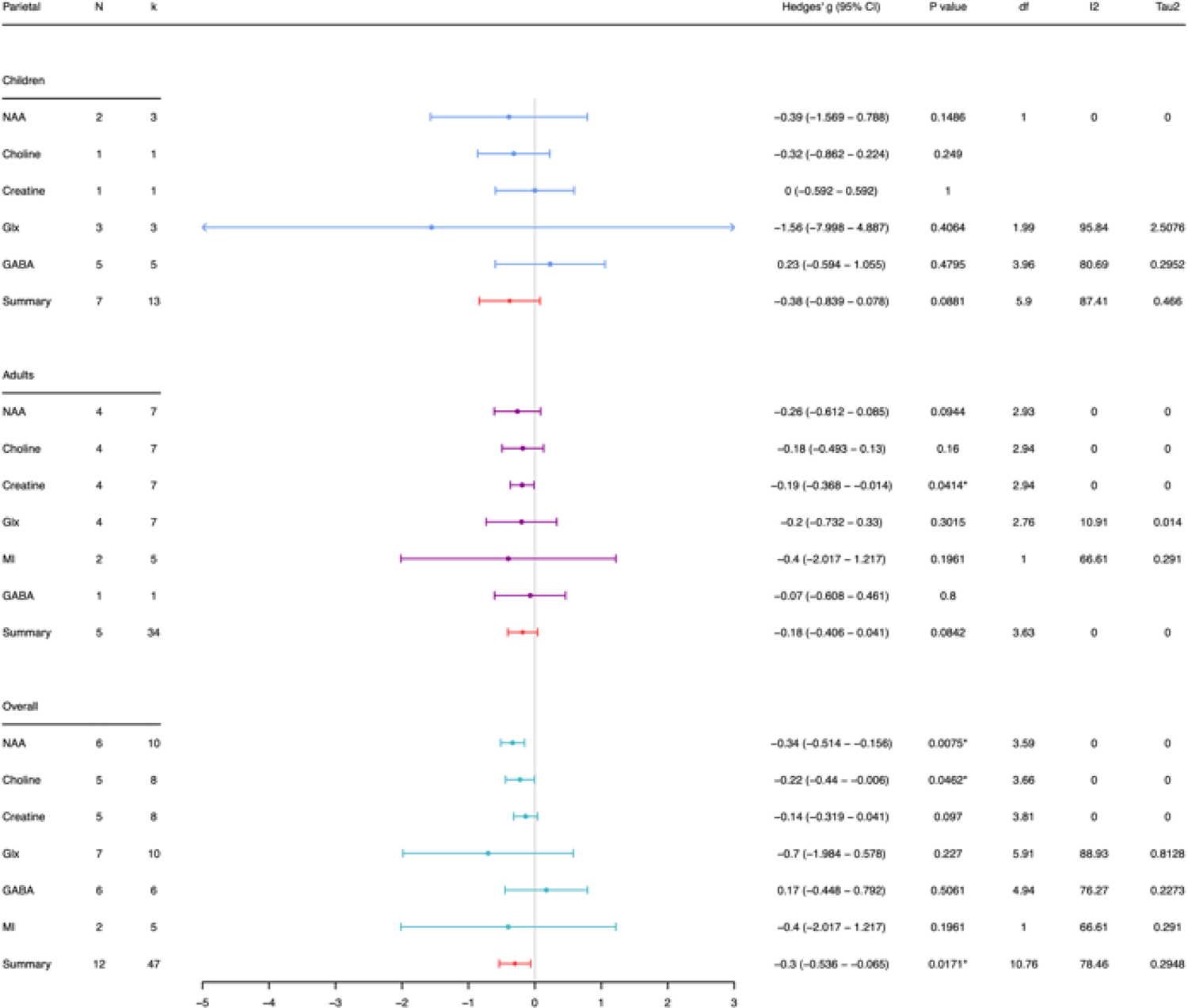
Summary forest plot findings in the parietal region split by metabolite and age group (children, adults, and pooled ages (overall)). N: number of studies, k: number of observations, % children: percentage of studies observing children only, I^2^: measure of between study heterogeneity, Tau^2^: Variance in true effect sizes (another measure of between study heterogeneity). Hedge’s g is reported in respect to autism. *Statistically significant at p < 0.05, and at p < 0.01 when the degrees of freedom < 4 for RVE t-tests.

**Figure 12.**
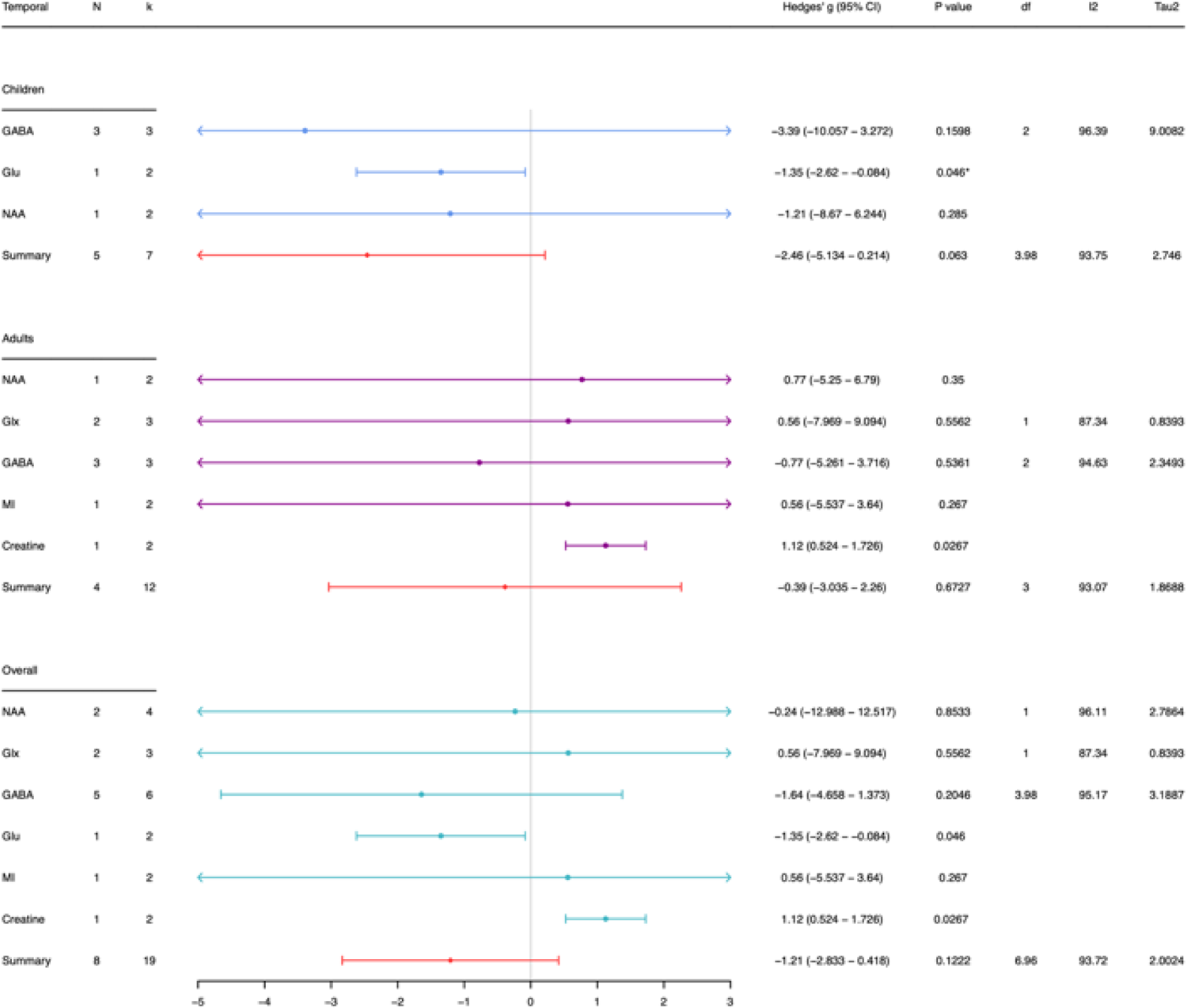
Summary forest plot findings in the temporal region split by metabolite and age group (children, adults, and pooled ages (overall)). N: number of studies, k: number of observations, % children: percentage of studies observing children only, I^2^: measure of between study heterogeneity, Tau^2^: Variance in true effect sizes (another measure of between study heterogeneity). Hedge’s g is reported in respect to autism. *Statistically significant at p < 0.05, and at p < 0.01 when the degrees of freedom < 4 for RVE t-tests.

To explore significant limbic findings in more detail, we grouped data by brain region grouping 2 (where limbic regions are differentiated) and age group. Again, the ACC was the only limbic region with sufficient study numbers to explore age effects. We observed a significant negative effect size for ACC GABA concentrations in children (Hedges’ g = −0.5, 95% CI: −0.67 – 0.32, I^2^ = 0, n = 4, p < 0.01; supplementary figure 3). In adults the study number was 1 (Hedges’ g = −0.21, 95% CI: −0.64 − 0.22, I^2^ = 0, n = 1, p = 0.34)..

**Figure 13.**
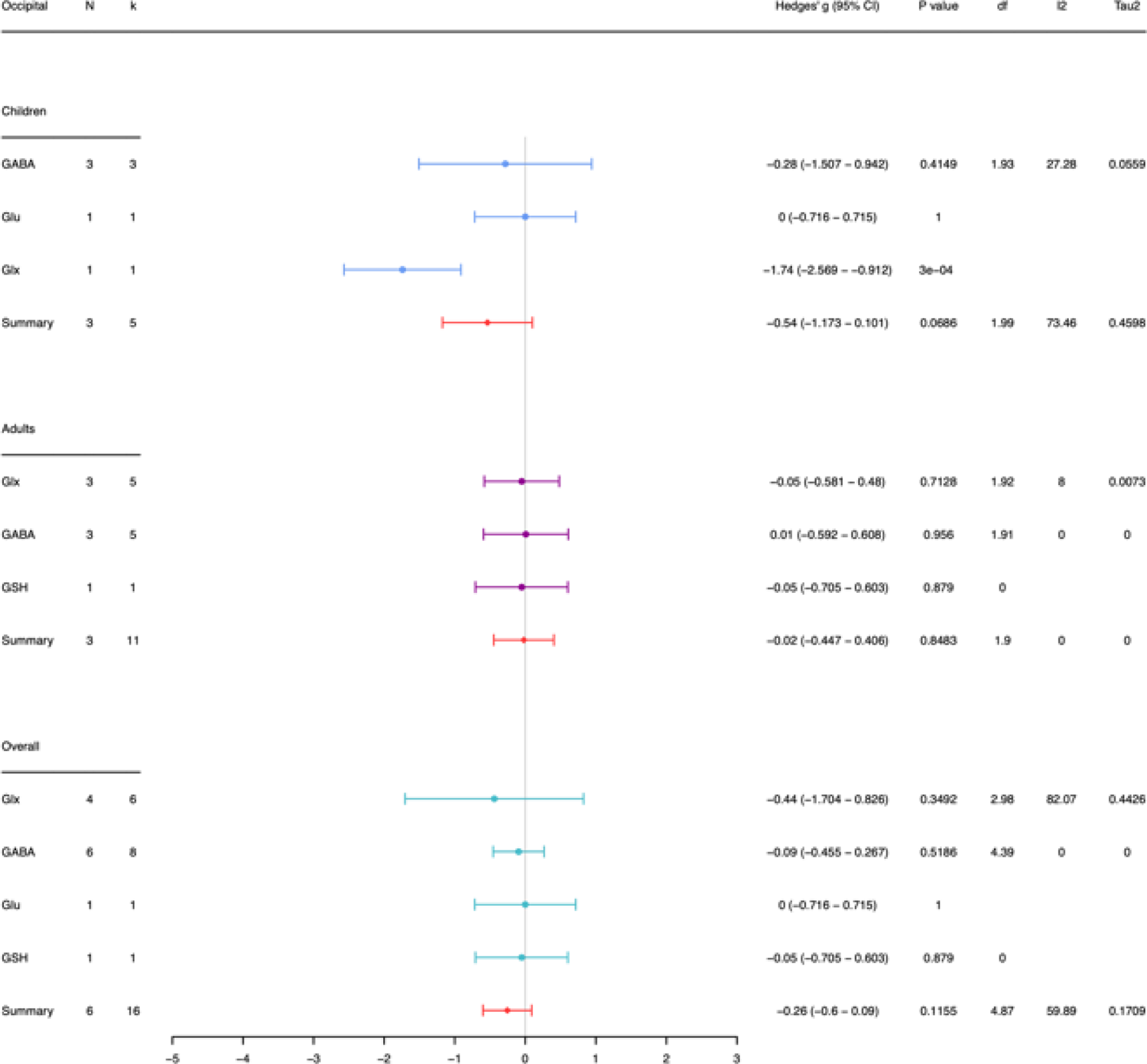
Summary forest plot findings in the occipital region split by metabolite and age group (children, adults, and pooled ages (overall)). N: number of studies, k: number of observations, % children: percentage of studies observing children only, I^2^: measure of between study heterogeneity, Tau^2^: Variance in true effect sizes (another measure of between study heterogeneity). Hedge’s g is reported in respect to autism. *Statistically significant at p < 0.05, and at p < 0.01 when the degrees of freedom < 4 for RVE t-tests.

#### Choline

*By brain region* A significant negative group effect size was found for studies measuring choline in the **parietal** regions (Hedges’ g = −0.22, 95% CI: −0.44 − −0.006, I^2^ = 0, n = 5, p < 0.05, 25.7% children; Figure 5). No significant effect size was observed for choline in the limbic (n=24; 44.1% children), cerebellum (n = 7; 75% children), frontal (n=1; 100% children), and prefrontal (n=9; 46.7 % children).

*By age group* We observed no significant effect size for studies measuring choline in adult only (Hedges’ g = 0.08, 95% CI: −0.12 − 0.28, I^2^ = 13, n = 15, p = 0.40; Figure 6) or children only subgroups (Hedges’ g = 0.06, 95% CI: −0.14 − 0.27, I^2^ = 44, n = 13, p = 0.50; Figure 6).

*By age group and brain region* When studies measuring choline where split by brain region and age group (Figure 7 − 13), no significant effect size was observed for any of the brain regions studied: cerebellum (n_children_ =5, n_adult_ =2; Figure 8), frontal (n_children_ =1, n_adult_ =0; Figure 8), limbic (n_children_ =12, n_adult_ =13; Figure 9), prefrontal (n_children_ =3, n_adult_ =6; Figure 10) or parietal (n_children_ =2, n_adult_ =4; Figure 11).

#### Creatine

*By brain region* A significant negative group effect size was observed for studies measuring creatine in the cerebellum (Hedges’ g = −0.23, 95% CI: −0.47 − −0.039, I^2^ = 0, n = 4, p < 0.05, 50% children; Figure 6). No significant effect size was observed for creatine in the limbic (n = 21; 46.4 % children), frontal (n = 1; 100% children), prefrontal (n = 10; 41.7 % children) and parietal regions (n = 6; 12.5 % children). For the temporal region n = 1.

*By age group* No significant effect size was observed for studies measuring creatine in adult only (Hedges’ g = 0.03, 95% CI: −0.23 − 0.28, I^2^ = 68, n = 15, p = 0.82; Figure 6) or children only subgroups (Hedges’ g = −0.38, 95% CI: −0.84 − 0.08, I^2^ = 79, n = 13, p = 0.097; Figure 6).

*By age group and brain region* When studies measuring creatine where split by brain region and age group (Figure 7 − 13), we observed a significant negative group effect size for **parietal** creatine in **adults** (Hedges’ g = −0.19, 95% CI: −0.37- −0.014, I^2^ = 0, n = 4, p <0.05; Figure 10) but not in **children** (Hedges’ g = 0.00, 95% CI: −0.59 − 0.59, I^2^ =NA, n = 2, p = 0.25; Figure 10).

No significant effect sizes were observed for the cerebellum (nchildren = 2, nadult = 2; Figure 7), frontal (n_children_ = 1, n_adult_ = 0; Figure 8), limbic (n_children_ = 11, n_adult_ = 11; Figure 9), prefrontal (n_children_ = 3, n_adult_ =7; Figure 10) or temporal regions (n_children_ = 0, n_adult_ = 1; Figure 12) in children or adults.

#### Glutamate and Glx

*By brain region* No significant effect size was observed for Glu in any region studied; limbic (n = 11; 40.7% children), cerebellum (n = 3; 66.7% children), frontal (n =2; 100% children), prefrontal (n = 2; 0% children). For the occipital and temporal regions n = 1.

Similarly, for Glx, no single brain region had a significant group effect size; limbic (n =19; 40.7% children), cerebellum (n = 3, 66.7% children), frontal (n = 3, 33.3% children), temporal (n = 2, 0% children), prefrontal (n = 7, 14.3% children), parietal (n = 7, 30% children) and occipital (n = 4, 16.7% children).

*By age group* No significant effect size was observed for studies measuring Glu in adults (Hedges G’ = −0.29, 95% CI: −0.78 − 0.21, I^2^ = 66, n = 4, p = 0.451; Figure 6) or children (Hedges’ g = 0.30, 95% CI: −0.031 − 0.63, I^2^ = 69, n = 9, p = 0.06; Figure 6). The same was observed for Glx (Hedges’ g_children_ = −0.27, 95% CI: −0.83 − 0.28, I^2^ = 84, n = 12, p = 0.30 and Hedges’ g_adults_ = −0.04, 95% CI: −0.18 − 0.25, I^2^ = 62, n = 17, p = 0.74).

*By age group and brain region* For Glu no significant effect size was observed for children or adults in any brain region studied: cerebellum (nchildren =2, nadult =1; Figure 7), frontal (nchildren =2, n_adult_ =0; Figure 8), limbic (n_children_ =8, n_adult_ =3; Figure 9), prefrontal (n_children_ =0, n_adult_ =2; Figure 10), temporal (n_children_ =1, n_adult_ =0; Figure 12) and occipital (n_children_ =1, n_adult_ =0; Figure 13).

The same was observed for Glx: cerebellum (n_children_ =2, n_adult_ = 1; Figure 7), frontal (n_children_ = 0, n_adult_ =1; Figure 8), limbic (n_children_ = 10, n_adult_ = 10; Figure 9), prefrontal (n_children_ = 1, n_adult_ = 7; Figure 10), parietal (n_children_ =3, n_adult_ = 4; Figure 11), temporal (n_children_ = 0, n_adult_ = 2; Figure 12) and occipital (n_children_ = 1, n_adult_ = 3; Figure 13).

#### GSH

*By brain region* When studies measuring GSH were split by brain region (Figure 5), no single brain region studied had a significant group effect size, this included limbic (n = 2, 0% children), frontal (n = 1, 0 % children), prefrontal (n = 2, 0% children) and occipital regions (n = 1, 0% children).

*By age group* GSH has only been studied in adult cohorts, for which no significant effect size was observed (Hedges’ g = 0.1, 95% CI: −1.28 − 1.48, I^2^ = 72, n = 3, p = 0.78).

#### mI

*By brain region* For studies measuring mI, no brain region studied had a significant group effect size (Figure 5): limbic (n = 16, 40% children), cerebellum (n = 3, 100% children), frontal (n = 3, 100% children), temporal (n = 1, 0% children), prefrontal (n = 4, 0% children), parietal regions (n = 2, 0% children).

*By age group* When studies measuring mI were split by age group (Figure 6), no significant effect size was observed for adults (Hedges G’ = −0.01, 95% CI: −0.19 − 0.20, I^2^ = 40, n = 10, p = 0.93) or children (Hedges’ g = 0.09, 95% CI: −0.86 − 0.68, I^2^ = 88, n = 10, p = 0.80).

*By age group and brain region* When studies measuring mI where split by brain region and age group (Figure 7 – 13), we observed no significant effect size for any brain region in children or adults: cerebellum (n_children_ =0, n_adult_ =3; Figure 7), frontal (n_children_ =2, n_adult_ =0; Figure 8), limbic (n_children_ =9, n_adult_ =7; Figure 9), prefrontal (n_children_ =0, n_adult_ =4; Figure 10), parietal (n_children_ =0, n_adult_ =2; Figure 11) and temporal (n_children_ =1, n_adult_ =1; Figure 12).

### 3.6 Effect of demographic factors

The effect of MRS study demographics on effect size is presented below. When split by metabolite and brain region the number of studies per group was too small for meaningful conclusions, as such in most cases we observed the effect of these factors across all brain regions and ages and in most cases, metabolites to maximise power.

#### Sex

Data was grouped by cohort sex ratio. Overall, 43 case-control comparisons included females (of these 10 included at least a 1:3 female:male ratio, 7 included at least a 1:1 female:male ratio), 13 studies excluded females, and 5 failed to report participant sex. Note this total is greater than the number of studies, because in some cases studies included variable cohorts for different brain regions studied.

Across all brain regions, ages and metabolites no obvious differences in group effect size were observed between studies that included at least 1:1 female:male ratio in cohorts (Hedges’ g_50%included_ = −0.1, 95% CI: −0.35 − 0.15, I^2^ =39, n =7, p = 0.36) and those that did not (Hedges’ g_50%not_ = −0.11, 95% CI: −0.30 − 0.078, I^2^ =49, n =51, p = 0.24). For studies that included 1:3 female:male ratio in cohorts, an effect size close to 0 was observed (Hedges’ g_1:3included_ = −0.02, 95% CI: −0.19 − 0.24, I^2^ =71, n =21, p = 0.82; Figure 14). For studies that did not include at least a 1:3 ratio of female:males, a greater, negative effect size was observed. (Hedges’ g_1:3not_ = −0.21, 95% CI: −0.47 − 0.042, I^2^ =84, n =34, p = 0.098; Figure 14).

**Figure 14.**
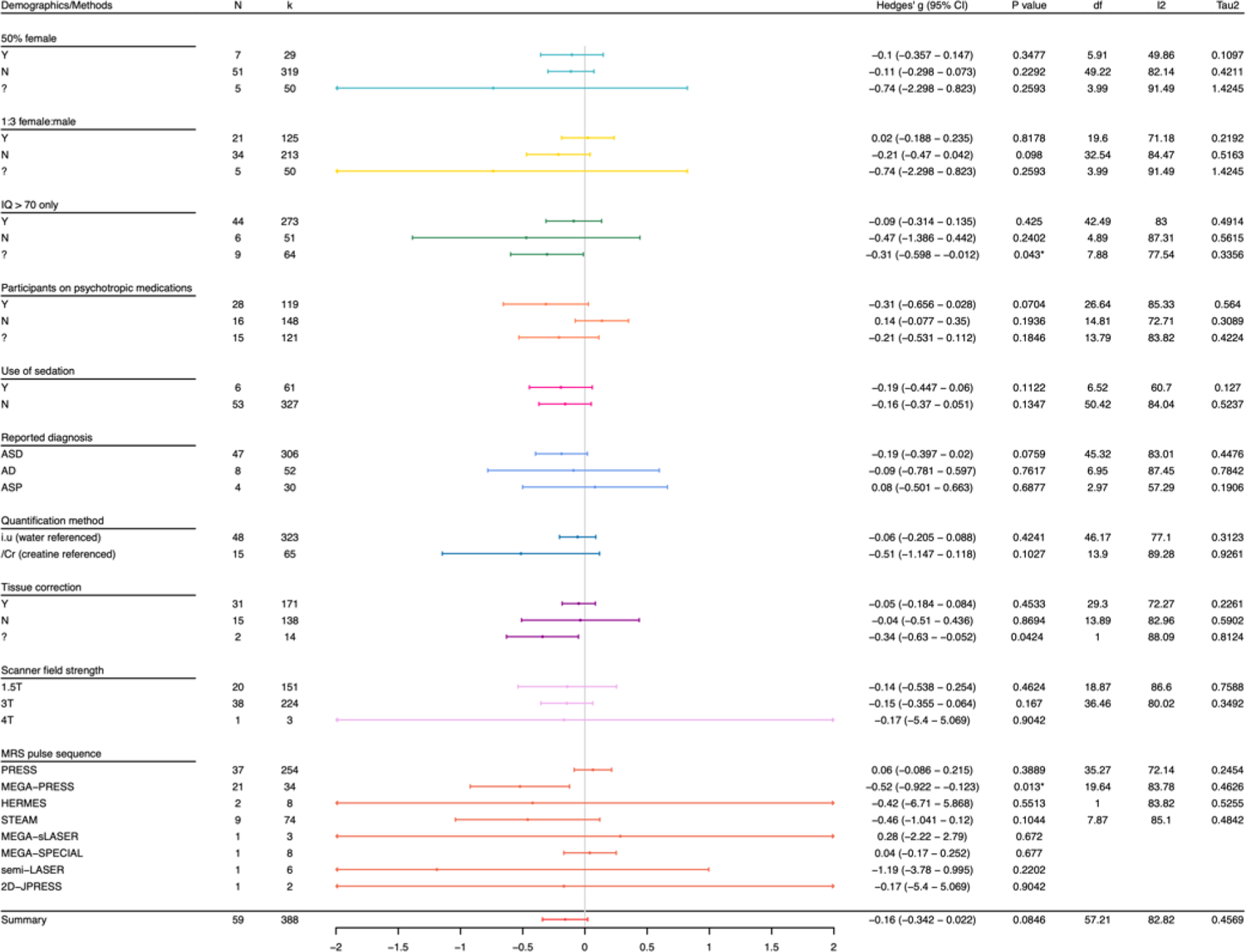
Forest plot summary of data grouped by demographic and MRS acquisition parameters. N: number of studies, k: number of observations, % children: percentage of studies observing children only, I^2^: measure of between study heterogeneity, Tau^2^: Variance in true effect sizes (another measure of between study heterogeneity). Hedge’s G is reported in respect to autism. *Statistically significant at p < 0.05, and at p < 0.01 when the degrees of freedom < 4 for RVE t-tests.

When data were grouped by metabolite, for studies measuring GABA (supplementary figure 4), those that included at least a 1:1 ratio of females to males in cohorts had a non-significant effect size close to zero (Hedges’ g = −0.03, 95% CI: −0.33 − 0.26, I^2^ = 0, n = 5, p = 0.76). Studies measuring GABA that did not include at least a 1:1 ratio of females to males in cohorts had a significant and negative effect size (Hedges’ g = −0.54, 95% CI: −1.00 − −0.082, I^2^ = 86, n = 19, p < 0.05). Note study numbers per group were too small to assess the effect of sex on limbic GABA in particular.

Studies measuring GABA that included at least a 1:3 female to male ratio in cohorts (supplementary figure 4) had a non-significant negative size (Hedges’ g = −0.25, 95% CI: −0.81 − 0.32, I^2^ = 80, n = 9, p = 0.35). Studies measuring GABA that did not include at least a 1:3 female to male ratio in cohorts had a significant and negative effect size (Hedges’ g = −0.59, 95% CI: −1.12 − −0.051, I^2^ = 85, n = 14, p < 0.05).

For studies measuring NAA (supplementary figure 5), those that included at least a 1:1 ratio of females to males in cohorts had a non-significant negative effect size (Hedges’ g = −0.55, 95% CI: −5.84 − 4.75, I^2^ = 86, n = 2, p = 0.42). Studies measuring NAA that did not include at least a 1:1 ratio of females to males in cohorts had a significant and negative effect size (Hedges’ g = −0.24, 95% CI: −0.42 − −0.054, I^2^ = 68, n = 30, p < 0.05).

Studies assessing NAA that included a minimum ratio of 1:3 females to males in cohorts (supplementary figure 5) had a non-significant negative effect size (Hedges’ g = −0.24, 95% CI: −0.65 − 0.16, I^2^ = 75, n = 11, p = 0.21). Studies measuring NAA that did not include at least a ratio of 1:3 females to males in cohorts had a significant and negative effect size (Hedges’ g = −0.24, 95% CI: −0.45 − −0.029, I^2^ = 67, n = 19, p < 0.05).

#### IQ

Data was grouped by cohort IQ. Overall, 44 case-control comparisons excluded individuals with IQ < 70, 6 studies did not exclude individuals with IQ < 70, 9 studies failed to report IQ of participants.

Across all metabolites, brain regions and age groups, studies that included individuals with an IQ < 70 had a non-significant negative effect size (Hedges’ g = −0.47, 95% CI: −1.39 − 0.44, I^2^ = 87, n = 6, p = 0.24; Figure 14). Those that excluded individuals with IQ < 70 had a non-significant effect size close to 0 (Hedges’ g = −0.09, 95% CI: −0.31 − 0.14, I^2^ =83, n = 44, p = 0.43 respectively; Figure 14).

When data were grouped by metabolite, for GABA (supplementary figure 4), studies excluding individuals with an IQ < 70 had a significant and negative effect size (Hedges’ g = − 0.27, 95% CI: −0.56 − −0.022, I^2^ = 74, n = 19, p < 0.05). Studies including individuals with an IQ < 70 had a non-significant negative effect size (Hedges’ g = −1.16, 95% CI: −4.36 − 2.05, I^2^ = 91, n = 3, p = 0.92). Note the study number discrepancy between these groups.

For studies measuring NAA (supplementary figure 5), those excluding individuals with an IQ < 70 had a non-significant and negative effect size (Hedges’ g = −0.23, 95% CI: −0.59 − 0.13, I^2^= 80, n = 22, p = 0.19). The same was observed for studies including individuals with an IQ < 70 (Hedges’ g = −0.37, 95% CI: −0.856 − 0.12, I^2^ = 73, n = 5, p = 0.10).

#### Psychotropic medication

Data were grouped by reported exclusion or inclusion based on psychoactive medication. 16 studies reported the exclusion of participants taking psychotropic medication at the time of scan. 28 did not exclude individuals on psychotropic medication at the time of scan. These studies reported varying proportions of participants on varying psychotropic medications, the most common being stimulants, anti-depressants, and anti-psychotics. 15 studies failed to report the medication status of participants.

Across all brain regions, ages and metabolites, studies excluding psychotropic mediation use had a non-significant positive group effect size (Hedges’ g = 0.14, 95% CI: −0.077 − 0.35, I^2^ = 72, n = 16, p = 0.18). Studies including individuals on psychotropic medications at time of scan had a non-significant negative group effect size (Hedges’ g = −0.31, 95% CI: −0.66 − 0.028, I^2^ = 85, n = 28, p = 0.070; Figure 14).

When data were grouped by metabolite, for GABA (supplementary figure 4), studies excluding psychotropic mediation use had a non-significant effect size at zero (Hedges’ g = 0.00, 95% CI: −0.40 − 0.42, I^2^ = 0, n = 3, p = 0.92). Studies that included individuals on psychotropic medications at the time of scan had a significant negative group effect size (Hedges’ g = −0.45, 95% CI: −0.84 − 0.050, I^2^ = 80, n = 18, p < 0.05).

For NAA (supplementary figure 5), studies excluding psychotropic mediation use had a non-significant negative effect size (Hedges’ g = −0.17, 95% CI: −0.46 − 0.12, I^2^ = 68, n = 12, p = 0.23). The same was observed for studies including individuals taking psychotropic medications (Hedges’ g = −0.45, 95% CI: −1.34 − 0.44, I^2^ = 87, n = 10, p = 0.28).

#### Sedation

Data was grouped by the reported use of sedation. In 6 studies, all or some of the participants received sedation during the MRI procedures: (Bejjani et al., 2012: 1 participant, intravenous propofol; Goji et al., 2017: all participants, 0.5mL/kg body weight of triclofos sodium; Harada et al., 2011: half of participants, 0.2 mL/kg body weight of triclofos sodium; Hashimoto et al., 1997: all children under 6, n = ?, triclofos sodium; Ito et al., 2017, all participants, 0.5mL/kg body weight of triclofos sodium; Mori et al., 2013: “most” participants, 0.5mL/kg body weight of triclofos sodium). 53 studies did not report use of sedation.

No obvious differences in group effect size was observed between studies that reported use of sedation (Hedges’ g = −0.19, 95% CI: −0.45 − 0.06, I^2^ = 60, n = 6, p = 0.11) and those that did not (Hedges’ g = −0.16, 95% CI: −0.37 − 0.051, I^2^ = 84, n = 53, p = 0.13; Figure 14; across all brain regions, ages and metabolites), although study number disparity between groups may contribute to this.

When data were grouped by metabolite, for GABA (supplementary figure 4), studies that reported use of sedation had a significant and negative effect size (Hedges’ g = −0.52, 95% CI: −0.80 − −0.25, I^2^ = 39, n = 4, p < 0.05). The same was observed for studies that did not report use of sedation (Hedges’ g = −0.44, 95% CI: −0.83 − −0.041, I^2^ = 83, n = 22, p < 0.05).

For studies measuring NAA (supplementary figure 5), studies that reported use of sedation had a negative effect size (Hedges’ g = −0.15, 95% CI: −0.59 − 0.29, I^2^ = 63, n = 7, p = 0.43).

Studies that did not report use of sedation had a significant and negative effect size (Hedges’ g = −0.38, 95% CI: −0.66 − −0.098, I^2^ = 79, n = 28, p < 0.05).

#### Reported diagnosis

Studies report various methods for autism classification of the included autism cohort, including classification according to the American Psychiatric Association’s Diagnostic and Statistical Manual, fourth addition (DSM-4; American Psychiatric Association, 2000), fifth addition (DSM-5; *Diagnostic and Statistical Manual of Mental Disorders*, 2013) and the International Classification of Diseases 10^th^ Revision (ICD-10, World Health Organization, 1993). Studies used various approaches for diagnosis including the Autism Diagnostic Interview—Revised ADI-R (Lord et al., 1994), the Autism Diagnostic Observation Schedule 1 (ADOS1; Gotham et al., 2007) and 2 (Lord et al., 2012); see Table 4 for specifics per study.

DSM-5 (American Psychiatric Association., 2013) criteria abandoned the sub-grouping of its processor DSM-4 (American Psychiatric Association., 1994), including autistic disorder (AD) and Aspergers syndrome (AS), under ‘ASD’ and facilitating the dual diagnosis of autism with other neurodevelopmental disorders such as ADHD (Colleen & Stone, 2014). As many MRS studies predate this change in classification, studies observing AD (as classified by DSM-4 and/or ICD-10), AS (as classified by DSM-4 and/or ICD-10), and ASD (as classified by DSM-5/ADI-R) were included in this meta-analysis.

Data were grouped by reported diagnosis/classification of autism cohorts (Table 4). 47 studies reported on a cohort classified as having ASD, inferred via the inclusion of multiple DSM-4/ICD-10 subgroups (AD & AS), and/or explicitly stating this classification in the study methods (DSM-5/ADI-R). Of note, 8 studies used DSM-4 classification but included individuals with AS or AD and so were classified as ‘ASD’ (see Table 4). 8 studies reported on individuals classified as having AD; this was inferred by the exclusion of any other developmental disorders (including AS) under the DSM-4 **and** the explicit statement of the inclusion of this classification only (DSM-4/ICD-10). 4 studies reported data from individuals with an AS classification, this being explicitly stated (DSM-4 and/or ICD-10).

Across all metabolites, brain regions and ages, studies reporting on ASD had a negative group effect size (Hedges’ g = −0.19, 95% CI: −0.40 − 0.02, I^2^ = 83, n = 47, p = 0.076; Figure 14). For studies reporting on AS, a group effect size close to 0 was observed (Hedges’ g = 0.08, 95% CI: −0.88 − 0.12, I^2^ = 57, n = 4, p = 0.69). The same was observed for studies reporting on AD (Hedges’ g = −0.09, 95% CI: −0.78 − 0.60, I^2^ = 87, n = 8, p = 0.76; Figure 14).

When data were grouped by metabolite, for GABA (supplementary figure 4), studies that reported on ASD had a significant and negative group effect size (Hedges’ g = −0.41, 95% CI: − 0.76 − −0.048, I^2^ = 80, n = 24, p < 0.05). Studies reporting on AS had a non-significant, negative group effect size (Hedges’ g = −0.33, 95% CI: −4.0 − 3.30, I^2^ = 37, n =2, p = 0.46). The same was observed for studies reporting on AD (Hedges’ g = −2.93, 95% CI: −26.62 − 20.77, I^2^ = 94, n = 2, p = 0.36).

For NAA (supplementary figure 5), studies that reported on ASD had a significant and negative effect size (Hedges’ g = −0.31, 95% CI: −0.61- −0.015, I^2^ = 78, n =24, p < 0.05). The same was observed for studies reporting AD (Hedges’ g = −0.61, 95% CI: −0.97 − −0.25, I^2^ = 70, n = 7, p < 0.05). For studies reporting AS, we observe a non-significant positive group effect size (Hedges’ g = 0.16, 95% CI: −0.49 − 0.82, I^2^ = 67, n =4, p = 0.48).

### 3.7 Effect of MRS methods

The effect of MRS parameters on effect size was also assessed.

#### Quantification methods & tissue correction

Data was grouped by metabolite quantification method: 48 studies reported metabolite concentrations in institutional units (i.u.; water scaled). 31 of these performed some form of tissue correction. 14 studies reported metabolite concentrations as creatine ratios only. In 3 studies, edited metabolite concentrations (GABA+/GSH) were recorded as creatine ratios, while unedited metabolites (NAA etc) were reported in institutional units (Cochran et al., 2015; Fung et al., 2021; Sapey-Triomphe et al., 2021).

Across all brain regions, ages and metabolites, the group effect size of studies reporting metabolite concentrations in institutional units was 0 (Hedges’ g = −0.06, 95% CI: −0.21 − 0.088, I^2^ = 77, n = 48, p = 0.42; Figure 14). The group effect size of studies reporting metabolite concentrations as creatine ratios was negative (Hedges’ g = −0.51, 95% CI: −1.15 − 0.12, I^2^ = 89, n = 15, p = 0.10; Figure 14).

When data were grouped by metabolite of interest (supplementary figure 5), studies reporting water-scaled NAA concentrations in i.u. had a significant negative group effect size (Hedges’ g = −0.29, 95% CI: −0.56 − −0.031, I^2^ = 77, n =27, p < 0.05). Studies reporting NAA concentrations as creatine ratios had a non-significant negative group effect size (Hedges G = −0.42, 95% CI: −0.57 − 0.12, I^2^ = 79, n =8, p = 0.11).

Studies reporting GABA concentration as a creatine ratio had a significant negative group effect size (Hedges’ g = −1.62, 95% CI: −2.97 − −0.27, I^2^ = 92, n = 7, p < 0.05; supplementary figure 4). Studies reporting water-scaled GABA concentration in i.u. had a non-significant negative group effect size (Hedges’ g = −0.11, 95% CI: −2.49 − 0.02, I^2^ = 37, n = 19, p = 0.09).

The group effect size of studies that reported the use of tissue correction (Hedges’ g = −0.05, 95% CI: −0.18 − 0.084, I^2^ = 72, n = 29, p = 0.45; Figure 14) was very similar to that of those that did not report the use of tissue correction for water-scaled data (Hedges’ g = −0.04, 95% CI: −0.51 − 0.44, I^2^ = 83, n = 15, p = 0.87; Figure 14).

#### Scanner field strength

Data was grouped by scanner field strength; 20 MRS studies were performed at 1.5 Tesla (T), 48 studies at 3T, and 1 study at 4T. No clear difference in group effect size was observed for the different scanner strengths (1.5, 3 & 4T; Figure 14; across all brain regions, metabolites, and ages).

When data were grouped by metabolite, for GABA (supplementary figure 4), one study at 1.5T had a non-significant negative effect size (Hedges’ g = −2.93, 95% CI: −26.62 − 20.77, I^2^ = 94, n = 1, p = 0.36). Most included studies measured GABA at 3T, for which a significant negative group effect size was observed (Hedges’ g = −0.40, 95% CI: −0.73 − −0.058, I^2^ = 80, n = 25, p < 0.05).

For studies that measured NAA at 1.5T (supplementary figure 5), we observe a significant negative effect size (Hedges’ g = −0.47, 95% CI: −0.91 − −0.037, I^2^ = 84, n = 19, p < 0.05). For studies measuring NAA at 3T, we observe a non-significant negative group effect size (Hedges’ g = −0.16, 95% CI: −0.37- 0.054, I^2^ = 52, n = 15, p = 0.13).

#### MRS sequence

Data was grouped by MRS acquisition pulse sequence used. 48 studies used non-editing MRS sequences; 37 studies used PRESS (Bottomley, 1987), 9 studies used Stimulated Echo Acquisition Mode (STEAM), 1 study used a two-dimensional J-resolved protocol (2D-JPRESS; (Joshi et al., 2013) and 1 study used semi-LASER (Pierce et al., 2021). 25 studies used editing MRS acquisition sequences; 21 studies used MEGA-PRESS; (Mescher et al., 1996, 1998), 2 studies used Hadamard Encoding and Reconstruction of MEGA-Edited Spectroscopy (HERMES; He et al., 2021; Saleh et al., 2016; Sapey-Triomphe et al., 2021) and 1 used MEGA-SPECIAL (Fung et al., 2021; M. Gu et al., 2018). Note 10 studies used both editing and non-editing sequences (Brix et al., 2015; Cochran et al., 2015; Drenthen et al., 2016; Harada et al., 2011; He et al., 2021; Ito et al., 2017; Maier et al., 2022; Pierce et al., 2021; Robertson et al., 2016; Wood-Downie et al., 2021).

Studies using MEGA-PRESS had significant negative group effect size (Hedges’ g = −0.52, 95% CI: −0.92 − −0.123, I^2^ = 83, n =17, p < 0.05; across all metabolites and brain regions; Figure 14). No significant effect size was observed for HERMES, STEAM, MEGA-SPECIAL, semi-LAZER, or JPRESS; Figure 14. Note 100% of studies using MEGA-PRESS were observing GABA.

When data were grouped by metabolite, for GABA (supplementary figure 4), studies that used MEGA-PRESS had a significant negative effect size (Hedges’ g = −0.52, 95% CI: −0.92 − − 0.12, I^2^ = 83, n =17, p < 0.05), which makes sense given the specificity of the MEGA-PRESS sequence. For GABA studies, as n = 1 for STEAM, MEGA-SPECIAL and HERMES acquisitions, results are excluded from interpretation.

For studies that used PRESS to measure NAA, a significant negative group effect size was observed (Hedges’ g = −0.24, 95% CI: −0.45- −0.024, I^2^ =74, n = 25, p < 0.05; supplementary figure 5). A non-significant group effect size was observed for studies measuring NAA using STEAM (Hedges’ g = −0.66, 95% CI: −1.46 − 0.13, I^2^ = 86, n = 9, p = 0.091). One study used MEGA-SPECIAL and one PROBE-P for NAA measurement (supplementary figure 5).

#### Voxel size and transients

Voxel size and the number of transients showed no relationship with study effect size across all metabolites (Spearman’s rho correlation = −0.019, p = 0.72 and Spearman’s rho correlation = 0.1, p = 0.0902 respectively; Figure 15).

**Figure 15.**
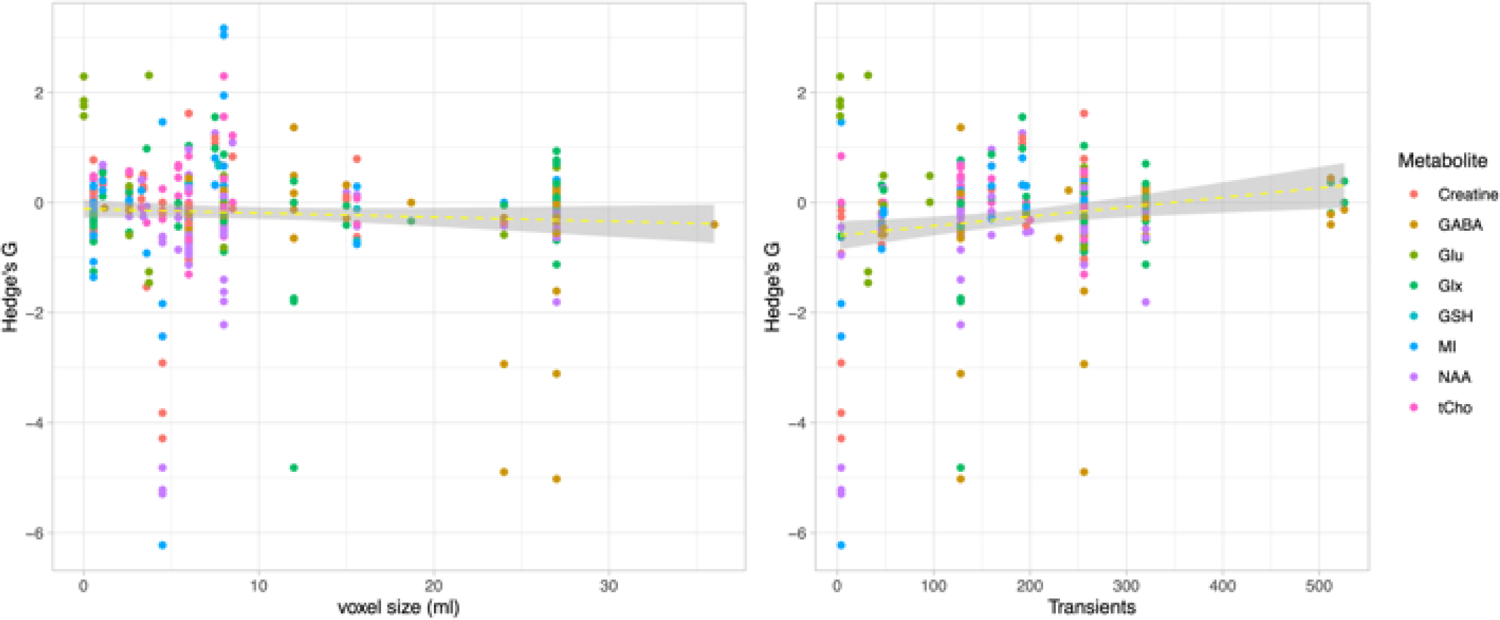
Relationship between study effect size and voxel size/number of transients. Voxel size (ml) showed no relationship with study effect size (Spearman’s rho correlation = − 0.019, p = 0.72), while the number of transients showed a positive correlation with study effect size (Spearman’s rho correlation = 0.1, p = 0.0902).

Data were split by metabolite and the correlation analysis was repeated. The number of transients had a significant positive correlation with effect size for NAA (Spearman’s rho correlation = 0.43, p = 0.000) and creatine (Spearman’s rho correlation = 0.47, p = 0.000), and mI (Spearman’s rho correlation = 0.42, p = 0.044). No significant correlation was observed between the number of transients and effect size for GABA (Spearman’s rho correlation = 0.22, p = 0.120), Glx (Spearman’s rho correlation = 0.14, p = 0.377), Glu (Spearman’s rho correlation = −0.43, p = 0.0534), GSH (Spearman’s rho correlation = −0.7, p = 0.122) and choline (Spearman’s rho correlation = −0.7, p = 0.122).

There was no significant correlation between voxel size and effect size for any metabolite; GABA (Spearman’s rho correlation = −0.24, p = 0.115), Glu (Spearman’s rho correlation = − 0.25, p = 0.214), GSH (Spearman’s rho correlation = −0.49, p = 0.325); Glx (Spearman’s rho correlation = 0.039, p = 0.771), NAA (Spearman’s rho correlation = 0.08, p = 0.476), choline (Spearman’s rho correlation = −0.018, p = 0.891), creatine (Spearman’s rho correlation = 0.081, p = 0.546) and mI (Spearman’s rho correlation = 0.14, p = 0.388).

### 3.9 Effect of MRS data quality

Analysis was repeated using only high-quality data as assessed by MRS −Q (Figure 16). General trends identified in the main meta-analysis were conserved, with small changes to effect size values but with no change in significance level. Across all brain regions and ages, significant negative group effect sizes were observed for NAA and GABA (Hedges’ g_NAA_ = − 0.23, 95% CI: −0.42 − −0.038, I^2^ = 64, n =23, p < 0.05 and Hedges’ g_GABA_ = −0.34, 95% CI: −0.62 − −0.059, I^2^ = 77, n = 25, p < 0.05).

**Figure 16.**
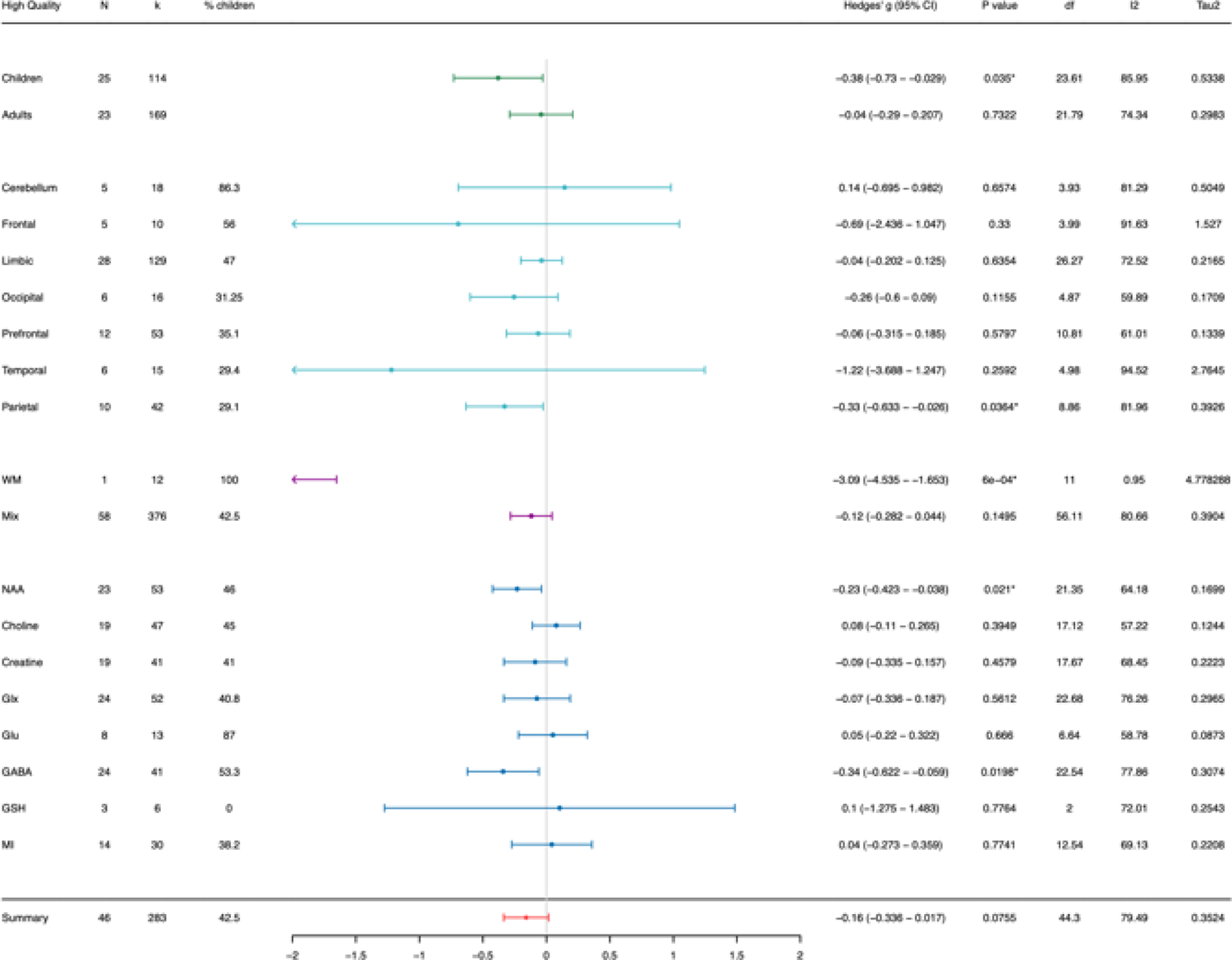
Data from only ‘high quality’ studies as assessed by MRS-Q, grouped by age (across all metabolites, brain regionsand ages), brain regions (across all metabolites and ages), voxel tissue type (across all metabolites and ages), and metabolite of interest (across all brain regions, ages, and tissue types). N: number of studies, k: number of observations, % children: percentage of studies observing children only, I^2^: measure of between study heterogeneity, Tau^2^: Variance in true effect sizes (another measure of between study heterogeneity). *Statistically significant at p < 0.05, and at p < 0.01 when the degrees of freedom < 4.

## 4. Discussion

### Summary of findings across all regions and ages

Here we provide evidence of significant alterations in brain metabolite concentrations in autism. Firstly, across all metabolites, we observe significantly lower metabolite concentrations in autistic children compared to non-autistic controls, whilst we observe no significant neurochemical differences between groups in adults. These findings support other MRI work suggesting that autistic brain differences are more pronounced in childhood compared to adulthood, potentially due to compensatory mechanisms (Courchesne, 2004; Schumann et al., 2004). However, we do find that these results are driven by changes in specific metabolites/brain regions which we explore at a mechanistic level below.

### Autism-associated alterations in NAA

We show that limbic and prefrontal NAA concentrations are significantly lower in autistic children compared to non-autistic controls. Of the limbic regions, this trend was potentially driven by the **anterior cingulate cortex (ACC),** however as this region was considerably more statistically powered than other limbic areas, this should be interpreted with caution.

Autism-associated differences in ACC and prefrontal NAA concentrations may indiciate differences in neuronal integrity within these brain regions, NAA being highly concentrated within the neuronal cell body (Rae, 2014). Consistent with this, the immature autistic brain displays global and regional structural differences when compared to non-autistic controls (Courchesne, 2004; Courchesne et al., 2001; Hazlett et al., 2005), including reductions in ACC and prefrontal GM and WM volumes as documented by various studies (Chien et al., 2021; Haznedar et al., 1997; Knaus et al., 2012; Yang et al., 2016), as well as reductions in ACC neuronal size and dendritic spine complexity (Kemper & Bauman, 1993; Simms et al., 2009). Lower NAA concentrations in childhood may also indicate untypicalities in developmental myelination, with decreased NAA concentrations in oligodendrocytes previously associated with myelin sheath lipid content abnormalities (Singhal et al., 2017). This is consistent with autism-associated reductions in the fractional anisotropy of ACC and prefrontal white matter tracts (indicative of demyelination and/or reduced nerve fibre coherence<u>; Barnea-Goraly et al., 2004; Basser, 1995; Beaulieu, 2002; Hau et al., 2019; Keller</u> <u>et al., 2007)</u> and post-mortem work finding autism associated reductions in prefrontal axon myelin thickness (Zikopoulos & Barbas, 2010). Finally, as NAA is synthesised in mitochondria (Siddiqui et al., 2016), differences in mitochondrial function in these regions may also contribute, with mitochondrial dysfunction recently linked to many autism aetiologies (see review; Siddiqui et al., 2016). These are not discrete factors; atypical mitochondrial function alters neuronal differentiation and connectivity (various models: Fernandez et al., 2019; Khacho et al., 2017; Klein Gunnewiek et al., 2020) and as such may contribute to the differences in neuronal integrity during brain development (Fernandez et al., 2019; Siddiqui et al., 2016).

These structural and cytoarchitectural differences may underlie observed differences in the functional activity and connectivity of both the prefrontal cortex (Cole et al., 2019; Gilbert et al., 2008; N. Schmitz et al., 2006; Woodward et al., 2017) and limbic brain regions (Etherton et al., 2011; Guo et al., 2019; Kana et al., 2007; Turner et al., 2006; Woodward et al., 2017; Y. Zhou et al., 2016) in autism. In turn, functional differences in these regions likely underpin behavioural and cognitive differences in autism, as for example the prefrontal cortex is associated with social cognition, executive function and language (Franklin et al., 2017; Friedman & Robbins, 2022; Mohapatra & Wagner, 2023; Sumiya et al., 2020), while limbic brain regions are involved in several brain processes relevant to autism phenotypes. The ACC, for example, has roles in attention and social cognition (Apps et al., 2016; Caruana et al., 2017; Chien et al., 2021; Dichter et al., 2009). The amygdala has been associated with facial recognition, facial expression processing, and emotional learning (Adolphs et al., 1994; Adolphs, 2008; Rutishauser et al., 2015; Wang et al., 2017). The thalamic nuclei relay primary sensory and motor information from the periphery to the appropriate cortical regions (Lopez-Bendito, 2018), while the hippocampus has been implicated in episodic memory (Scoville & Milner, 1957; Eldridge et al., 2005) and more recently social interaction (Tavares et al., 2015; Banker et al., 2021). As such, researchers find a negative correlation between limbic (ACC) and prefrontal NAA concentrations and autism clinical scores (incl. communication impairments, IQ, autistic mannerisms & sensory processing abnormalities; Gabis et al., 2008; Hegarty et al, 2017; Carvalho Pereira et al., 2017; Brix et al., 2015; Edmondson et al., 2020). NAA differences are therefore mechanistically relevant as, in the context of the wider structural and functional implications, they contribute to (and may help to predict) autism phenotypes and their severity (Denisova & Lin, 2023).

By adulthood, there is a convergence of limbic and prefrontal NAA concentrations between autism and non-autistic controls, with significant differences between the two groups in prefrontal and limbic (ACC) NAA concentrations no longer seen, this finding unbiased by statistical power (same number of studies per age group). As discussed, this is not the first time a ‘normalisation’ of autism brain differences has been identified in the MRI imaging field; significant atypicality’s in brain structure observed during early childhood have been found to be absent in adult cohorts, potentially explaining the loss of NAA differences (Courchesne, 2004; Courchesne et al., 2001; Hazlett et al., 2005). However, this is largely based on cross sectional data (as is our meta-analysis) and is in conflict with other MRI and human post mortem findings that show cytoarchitectural differences at the neuronal and regional level persist in adulthood (Kemper & Bauman, 1993; Simms et al., 2009; Yang et al., 2016). As such, in the absence of longitudinal evidence, a ‘normalisation’ hypothesis should be considered with caution (Yankowitz et al., 2020).

### Autism-associated differences in GABA and Glutamate (Glu and Glx)

We also find autism-associated differences in GABA concentrations, with **limbic** GABA concentrations significantly lower in autistic compared to non-autistic children. This trend is notably driven by the **ACC** within the limbic regions studied. For autistic adults, we find no significant alterations in GABA concentrations. However, due to the limited number of studies focusing on GABA concentrations in adults, particularly in the ACC/limbic regions, an observation bias may underpin this apparent age effect.

The MRS GABA+ signal is a proxy measure for the degree a circuit can employ inhibition (Rae et al., 2014; Puts et al., 2015). In this context, the observed alterations in GABA concentrations (given that we observe no significant alterations in Glx/Glu concentrations) are consistent with the E/I imbalance theory of autism (Rubenstein & Merzenich, 2003), specifically showing an uncompensated shift in E/I balance towards excitation. While the relatively poor spatial resolution of the MRS technique means we cannot specify the cellular mechanisms that underpin these observations, previous work observes autism-associated reductions in the expression of the enzymes responsible for GABA synthesis (glutamate decarboxylase 65 and 67; Fatemi et al., 2002), as well as autism-associated decreases in parvalbumin (PV+) interneuron density, the primary cortical inhibitory (and so GABA expressing) interneuron (Hashemi et al., 2017). Both could sensibly contribute to the observed differences in limbic (ACC) GABA concentrations.

At the mechanistic level, local GABA concentrations have previously been associated with functional magnetic resonance imaging (fMRI) task response amplitudes (Muthukumaraswamy et al., 2009; Sampaio-Baptista et al., 2015; T. W. Schmitz et al., 2017) and due to GABAergic regulation of neuronal synchrony within local and regional brain circuits (Baumgarten et al., 2016; Muthukumaraswamy et al., 2009), the functional connectivity strength of various networks (Muthukumaraswamy et al., 2009; Stagg et al., 2014; Assaf et al., 2010; Sampaio-Baptista et al., 2015; Y. Zhou et al., 2016). Both ACC functional connectivity and task-induced activation have been shown to be atypical in autism (Agam et al., 2010; Assaf et al., 2010; Cole et al., 2019; Kana et al., 2007; Y. Zhou et al., 2016) and have been associated with repetitive behaviours and social differences (Chien et al., 2021; Thakkar et al., 2008; Yang et al., 2016). This provides a mechanistic link between differences in limbic (ACC) GABAergic activity and autism phenotypes, with previous work finding negative correlations between ACC GABA concentrations and autism clinical scores (Carvalho Pereira et al., 2017; Edmondson et al., 2020; Brix et al., 2015). Our results may thus help to explain why pharmacological potentiators of the GABAergic system improve behavioural and cognitive phenotypes in mouse models of autism (Baclofen; Silverman et al., 2015, Gaboxadol; Cogram et al., 2019) and in autistic children (Bumetanide; Lemonnier et al., 2012, 2017; Mahdavinasab et al., 2019; Zhang et al., 2020). However, according to our results, shifts in E/I balance may not be global or equal across the brain, whilst GABAergic drugs act without spatial specificity and so may have (counter-productive) off target effects. This potentially explains the null findings during phase 2 autism clinical trials for Arbaclofen, a GABA-b receptor agonist (and so potentiator; Veenstra-VanderWeele et al., 2017), as well as the sometimes moderate side effects reported with other agents (Bumetanide; Zhang et al., 2020).

There are several limitations specific to the measurement of GABA and Glu/Glx that require some discussion. First, most of studies included in this meta-analysis measured GABA concentrations only, yet decreases in GABA may lead to compensatory decreases in Glu (Kubas et al., 2012). While we observe no significant differences in Glu/Glx concentrations across all brain regions and ages (Glu/Glx observations were of a similar number to GABA), to resolve this definitively, multiple measurements should be taken within the same cohort or even at the individual level. Furthermore, care must be taken to equate MRS-measured GABA and Glu to neurotransmission E/I balance given their role in energy metabolism (Rae 2014). These metabolites are in constant flu as GABA (and GSH) are synthesised downstream of Glutamate/Glutamine. As such, MRS GABA/Glu balance may only be an indicator of broader tonic E/I in the brain.

Secondly, roughly half of studies measuring GABA reported concentrations as GABA plus macromolecule (MM) contamination of signal (GABA+), while the remaining studies used MM modelling to obtain pure GABA estimates. While we did not report on the effect of this on study outcome (as modelling approaches varied between studies and thus were likely not comparable) and the role of MM in autism is likely to be relatively small, it does represent another source of potential bias. Although MM-suppressed editing sequences are available, they are particularly sensitive to motion due to subtraction of non-interlaced transients (Edden et al., 2016) and as such may not be optimal in populations who tend to move more in the scanner environment such as those with cognitive difficulties or autism (Yerys et al., 2009, Puts et al., 2017).

Lastly, we recorded Glu and Glx separately and findings were consistent. However, it remains unclear whether Glu can be measured precisely at 1.5T and 3T and can be measured separately from Glx (glutamate + glutamine). In addition, quantification of Glx will differ between studies, some having quantified this from unedited MRS, others from the Glx peak in the GABA-edited difference spectrum, and others from the SUM (edit-ON + edit-OFF) or edit-OFF spectrum. It is well-known that measures derived using these different approaches do not always equate to one another (Bell et al., 2021). While Glu appears to contribute in majority to MRS Glx (as opposed to Gln; F. Du et al., 2009; Tapper et al., 2020), further caution should be taken interpreting measurements in the context of E/I balance.

### Choline, Creatine and mI

The number of studies (and percentage of child cohorts) observing creatine, choline and mI was not dramatically smaller than those observing GABA and NAA. As such, we do not think that our lack of significant findings for these metabolites is due to a bias in the total number of observations. There is however some evidence of autism-associated differences in these metabolites. We find significantly lower parietal choline (and NAA) concentrations and cerebellar creatine concentrations in autism. These changes were not significant in either the children only or adults only subgroups (only when combined across all ages), likely due to a lack of power from the smaller study numbers.

MRS total choline is composed of the choline containing compounds phosphocholine, phosphatidylcholine and glycerophosphocholine (Miller et al., 1996; Rae, 2014), which together are essential for neuronal membrane integrity (Rae, 2014). Reduced parietal total choline (and NAA) may therefore be indicative of previously characterised structural differences in this region, including reductions in GM and parietal cortical thickness (Ford & Crewther, 2016b; McAlonan et al., 2005; Wallace et al., 2010). MRS creatine signal is composed of creatine, which when reported as total creatine includes its phosphorylated form phosphocreatine, which acts as a readily available and fast energy store within neurons (Rae, 2014; Wallimann et al., 1992). The cerebellum is widely understudied in autism, however differences in cerebellar creatine may reflect abnormalities in neuronal activity within this region, in keeping with previously described alterations in cerebral-cerebellar functional connectivity and activity in autistic individuals (Ford & Crewther, 2016a; Mapelli et al., 2022; Mostofsky et al., 2009).

### Redox hypothesis (GSH)

Recent advances in MRS acquisition sequences have facilitated the quantification of GSH *in viv*o (Terpstra et al., 2003) through spectral editing, and so may provide evidence to support the redox hypothesis. Of the six relevant studies, no significant differences in GSH concentrations between autistic adults and non-autistic controls were identified (Limbic regions, frontal regions and occipital regions). If GSH conforms with the general trend identified in this meta-analysis, future work may find more significant GSH differences in childhood.

### Effect of MRS study design

Demographic and methodological factors likely explain why between study heterogeneity (I^2^) was large, even when data were simultaneously grouped by metabolite, age and brain region. As such, we explored how these factors associated with effect size.

### Sex

Males are diagnosed with autism more often than females, with a reported 1:3 ratio of females to males in autistic populations (Loomes et al., 2017). Recent work suggests that the presentation of autism in females may simply not be captured with current gold standard tools used by researchers to confirm autism diagnosis, leading to underestimates of female autism prevalence and the exclusion of females from research studies at significantly greater rates than males (de Giambattista et al., 2021; D’Mello et al., 2022). In this meta-analysis, only 16% of studies included cohorts matching population prevelance (1:3 female:male), only 11% included sex-matched cohorts (1:1 female:male), while 22% of included studies excluded females altogether. Failure to include sex matched samples diminishes statistical power for sex comparison, which is vital as the underlying biochemistry of autism may differ between sexes (Fung et al., 2021). As an example, we observed lower concentrations of NAA and GABA in autism *only* when female:male ratios of cohorts were less than 1:3/1:1. Thus we cannot be certain that the main outcomes of this meta-analysis are not specifically biased for male populations, with potentially subtler metabolite differences in autistic females. Future MRS study design should thus enable reliable statistical evaluation of sex differences to address this.

### IQ

75% of studies included in this meta-analysis excluded individuals with an IQ < 70, although it is estimated that around 30% of autistic children have an intellectual disability (Christensen et al., 2016; Zablotsky, 2017). As such, our results are more applicable to individuals without intellectual disability (i.e., having low support needs). Across all metabolites, our findings show tendencies that neurometabolic alterations in individuals with IQ > 70 are subtler than those with more serve intellectual impairments. This agrees with studies showing that altered neurometabolite levels (specifically NAA and GABA) linearly correlate with IQ (Carvalho Pereira et al., 2018; Gabis et al., 2008; Hegarty et al., 2018). While IQ should thus be considered as a potentially confounding factor in MRS studies of autism, discounting those with intellectual disabilities reduces the generalizability of study results, an important consideration as those most likely to be eligible/desire therapeutic intervention are those with higher support needs. Thus, excluding those with IQ < 70 with the sole aim of controlling for IQ is not necessarily an advantageous or suitable approach if justifying the same study with therapeutic goals.

### Sedation and psychotropics drugs

According to a study in 2017, 50% of autistic children take psychotropic medication (Madden et al., 2017). Here, 47% of included studies contained participants using psychotropic medications (n = 28), with SSRI’s being the most common. Psychotropic medication use by autistic individuals may indicate more severe symptom profiles, as well as the treatment of co-occurring psychiatric conditions such as anxiety disorder, ADHD and depression, which are very common in autism (72% of autistic individuals; Strang et al., 2012; Hours et al., 2022)) and also associated with alterations in the GABAergic system (Edden et al., 2012; Hasler et al., 2007; Pollack et al., 2008). Thus, the direct impact of psychotropic medications on brain metabolites (Sanacora et al., 2002) in addition to the presence of co-occurring disorders and/or an increased phenotype severity may contribute to our findings that GABA alterations are more pronounced in studies including participants on psychotropic medication compared to studies excluding such participants. As only 24% of the studies included in this meta-analysis controlled (statistically) for psychotropic medication usage in their analysis and only half reported co-occurring conditions. In future, study designs should enable statistical comparison of medicated and non-medicated groups, and transparent reporting of co-occurring disorders is recommended.

10% of the included studies used sedation during MRI procedures, most commonly using triclofos sodium. Across studies, there was considerable variability of both the dose and proportion of sedated participants (from less than half – all). Based on these studies alone, we observed no clear effect of sedation on study outcome. While the impact of triclofos sodium on brain metabolites has not been systematically assessed, these findings are consistent with a number of studies that find no impact of sedation or anaesthesia on brain metabolite NAA, creatine and Glu concentrations (F. Du et al., 2009; pentobarbital; Jacob et al., 2012; Sevoflurane and Propofol). Note however that the effect of anaesthesia on GABA levels in particular is undefined.

### Diagnosis

In line with current DSM-5 guidelines, studies observing AD, AS, and ASD were included in this meta-analysis. Nevertheless, across all metabolites, we observed trends for greater metabolite differences in ASD cohorts compared to those classified to either an AS and/or AD. More specifically, significantly lower GABA concentrations were observed in ASD cohorts only, whilst significantly lower NAA concentrations were observed in AD and ASD cohorts only. As such, the main outcomes of this meta-analysis are at this stage applicable to ASD cohorts (reported by 72% of included studies), partially applicable to AD cohorts (reported by 13% of studies included) and not applicable to AS cohorts (reported by only 6% of studies included). As AS diagnosis historically included mainly high-functioning individuals with ‘milder’ phenotypes compared to AD (Colleen & Stone, 2014), it is perhaps not surprising that no significant metabolite differences are observed for this group, particularly given that metabolite alterations correlate with phenotype severity (see above). In future, exploring the neurochemistry of non-subjective clinical behaviours maybe a preferable approach to mitigate the confounding nature of evolving diagnostic criteria between studies.

### Quantification method

We observed trends for stronger autism-associated neurometabolite differences in creatine referenced data compared to water referenced. This is striking as differences in GABA concentrations were only significant when creatine referenced, yet 60% of studies reported water-referenced GABA concentrations. There are multiple factors that may contribute to these results. First, creatine referencing typically assumes that creatine does not differ in the autistic brain which may not be the case; as evidenced by the observed significant differences in cerebellum creatine concentrations between autism and non-autistic groups. On the other hand, water referencing requires corrections for tissue composition and metabolite relaxation to obtain estimated metabolite concentrations (as recommended by MRS-Q). Given the breadth of studies on anatomical differences in autism at both the micro- and meso-level (see review; (Donovan & Basson, 2017), this complicates partial volume correction and introduces various unknowns. Both methods thus have their limitations and so it would be beneficial to study the structural and neurochemical associations in autism in more detail. However, at present, it is suggested that researchers report both ratio and estimated concentrations for transparency purposes.

### Scanner field strength

Although we observed no effect of scanner field strength on study outcome overall, for NAA we observed significantly lower concentrations in autism at 1.5T and not 3T (although the effect size at 3T remained negative). Since 3T provides a higher SNR compared to 1.5T, this may be cause for concern (Di Costanzo et al., 2007; Wilson et al., 2019, 2019). However, NAA is one of the most highly concentrated metabolites on the MRS chemical shift spectra/axis, making it one of the most accurate to quantify at low field strengths (1.5T) (Peek et al., 2020). As such, we can be confident that the significance of findings at 1.5T is not just a result of extra noise (the same could not be said for low concentration metabolites for example Glx/GABA). Furthermore, the significance of NAA findings was conserved when low quality studies were excluded (discussed below), providing further reassurance that the observed data trends are not skewed/inflated by low quality data.

### Pulse sequence

The impact of MRS pulse sequence on study effect size is more challenging to asses since study number per sequence was often too small to comment (n = 1 for MEGA-sLASER,, MEGA-SPECIAL, semi-LASER, 2D-JPRESS). Across all metabolites, we observed significant alterations for studies using MEGA-PRESS editing sequence only. As 100% of studies using MEGA-PRESS observed GABA concentrations, this was likely driven by the observed reductions in GABA concentrations in autism as opposed to a bias introduced by the sequence itself. Lower NAA concentrations (in autism) were observed by studies using PRESS and STEAM non-editing sequences, while this was only significant for PRESS. Due to differences in sequence design, STEAM achieves more accurate volume selection (less chemical shift displacement) compared to PRESS, however at the expense of SNR, achieving around half the SNR of PRESS (van der Graaf, 2010). As such, perhaps with greater SNR (using PRESS) we can better detect NAA differences in autism. The discussion below however challenges this notion for NAA.

### Transients and voxel size

Increasing the number of transients and voxel size increases the accuracy of MRS metabolite quantification (due to increasing signal to nose ratio (SNR); Mikkelsen et al., 2018). Optimally, increasing accuracy should increase the ability to detect differences in metabolite concentrations between groups if present (as suggested above). As such it was surprising that overall, we observed no significant interaction between voxel size/transients and study effect size. In fact, for NAA, we found a significant positive correlation between the number of transients and study effect size. This suggests that with increasing accuracy studies can better detect increases in NAA concentrations in autism (hedges g is positive), and as such queries the validity of our main results. However, multiple MRS parameters (alongside transients) contribute to MRS data quality, as was comprehensively assessed by MRS-Q. MRS-Q is thus perhaps a better assessment of whether the observed data trends are reliable than looking at the number of transients alone.

### Publication bias

For studies observing GABA and NAA we observed a significant publication bias, with study effect sizes positively skewed (absence of effect sizes in the positive direction). Note that all of the GABA studies and the majority of the NAA that studies were identified as driving this asymmetry (large, significant negative effect sizes) reported creatine scaled metabolite concentrations (Gabis et al., 2008; Gaetz et al., 2014; Kubas et al., 2012; Port et al., 2019), reiterating the potential bias quantification method introduces to study outcome.

Importantly however, between study heterogeneity was large and so included studies likely do not truly share one true effect size (due to real differences in demographics/methodology), making such bias assessments potentially inappropriate (Peters et al. 2007; Terrin et al. 2003; Simonsohn et al., 2014). As such the identified studies remained in our meta-analysis.

Biases out of the scope of a meta-analysis do exist, as identified in our AXIS assessment; the use of hospital controls instead of the general population (often migraine patients), failure to characterises individuals who failed to complete the MRS scan, failing to control for co-occurring conditions such as ADHD and anxiety in particular, and variation in individual researcher MRS processing pipelines (Bhogal et al., 2017) all likely affect these data.

Elimination of all confounding factors in autism case-control studies, this being a highly heterogenous and complex group, is perhaps unrealistic. Better and transparent reporting practises would make statistical control for this bias easier and interpretation of results more reliable.

### Effect of MRS quality on study outcome

The development of consensus guidelines (MRSinMRS and MRS-Q; A. Lin et al., 2021; Peek et al., 2020) is essential for improvement of adequate reporting and acquisition of MRS data. Without this, it remains a considerable challenge to isolate non-confounded data trends as proven by this meta-analysis. This consensus was used to assess the quality of MRS study design and data. Low quality studies (10%) generally did not report sufficient scanner field strength, voxel size and transients (consensus edited; 3T and 27 ml voxel for edited MRS for GABA, 240 transients. Unedited; 3.4 ml voxel, 128 transients at 3T). As discussed, these parameters are associated with SNR and directly influence the quality of the acquired MRS spectra (Mikkelsen et al., 2018). To prevent the skewing of data trends due to poor quality data, we excluded these studies from analysis. All significant trends from the main meta-analysis were conserved; significantly lower NAA and GABA concentrations in autism compared to TD. As such, we present high-quality evidence of autism-associated alterations for NAA and GABA concentrations in limbic (and parietal) regions. While MRS-Q addresses MRS methodological factors, these results are still subject to the demographic confounds discussed above.

### Limitations of the current meta-analysis

One limitation to this meta-analysis is that we initially included all studies regardless of quality as assessed by MRS-Q. However, we observed no changes to the data trends when low quality data were excluded, and we used subgrouping to thoroughly explore potential biases in study outcomes. We have also been very transparent in reporting of these (MRS study design and demographics). For some brain regions we lacked statistical power due to the small number of studies (particularly the temporal and occipital regions), and as such the lack of significant differences in neurometabolite concentrations between groups should be interpreted with caution as it may simply reflect that we are unable to detect differences at this time.

A key strength of the current analysis is that we employed a conservative RVE method for Hedges’ g calculation (Pustejovsky & Tipton, 2022), this prevented the skewing of outcomes by dependent data, with most studies taking multiple metabolite measurements (and voxels) from one individual (Borenstein et al., 2021). This was not considered by previous systematic reviews (Ajram et al., 2019; Y. Du et al., 2023; Ford & Crewther, 2016a). We have also comprehensively assessed the impact of study design on outcome to expose confounding factors in our results.

## Conclusion

Overall, in this meta-analysis, we find evidence of strong autism-associated alterations in neuro-metabolite concentrations. More specifically, we find that autistic *children* have significantly lower concentrations of NAA in the limbic and prefrontal regions, perhaps indicative of structural differences and/or mitochondrial disfunction in this region. Also reported was evidence of uncompensated shifts in E/I imbalance; autistic *children* have significantly lower GABA concentrations in limbic regions (more specifically the ACC), while we observe no significant alteration in Glx/glutamate concentrations. The severity of alterations in both NAA and GABA has been shown to correlate with the severity of autism clinical scores, supporting the hypothesis that such atypicality’s contribute mechanistically to the autism phenotype, with GABA specifically representing a valid therapeutic target. Finally, we observed that sex, co-occurring disorders, psychotropic medication use and MRS metabolite quantification approach influence the degree of GABA/NAA alteration observed and so likely contribute to the heterogeneity of previous findings. Compliance with consensus MRS parameters (e.g MRS-Q), paired with appropriate and transparent reporting is essential to address this and improve the ease at which reliable trends can be identified and accepted in future.

## Data Availability

All data and analysis files are available

https://osf.io/uv67s/

## Acknowledgements

TA was supported by an MRC Clinician Scientist Fellowship [MR/P008712/1] and Transition Support Award [MR/V036874/1]. AT, NP and TA received support from the Medical Research Council Centre for Neurodevelopmental Disorders, King’s College London [MR/N026063/1]. The views expressed are those of the author(s) and not necessarily those of the NHS, the NIHR or the Department of Health and Social Care. The funders had no role in the design and conduct of the study; collection, management, analysis, and interpretation of the data; preparation, review, or approval of the manuscript; and decision to submit the manuscript for publication.

