## supplementary information for "Neurometabolite differences in Autism as assessed with Magnetic Resonance Spectroscopy: a systematic review and meta-analysis"

### 1. Supplementary

**Supplementary table 2:** Data extraction table.

Sd = standard deviation, IQR = inter quartile range.

| |
|---|
| Study |
| Voxel brain region |
| Group (control or autism) |
| NAA mean |
| NAA sd |
| NAA median |
| NAA IQR |
| NAA range |
| choline mean |
| choline sd |
| choline median |
| choline IQR |
| choline range |
| Pcr + Cr mean |
| Pcr + Cr sd |
| Pcr + Cr median |
| Pcr + Cr IQR |
| Pcr + Cr range |
| creatine mean |
| creatine SD |
| creatine median |
| creatine IQR |
| creatine range |
| Glx mean |
| Glx sd |
| Glx median |
| Glx IQR |
| Glx range |
| Glu mean |
| Glu sd |
| Glu median |
| Glu IQR |
| Glu range |
| Gln mean |
| Gln sd |
| GABA+ mean |
| GABA+ sd |
| GABA+ median |
| GABA+ IQR |
| GSH mean |
| GSH sd |
| mI mean |
| mI sd |
| mI median |

| |
|---|
| mI range |
| lac mean |
| lac sd |
| ASD n |
| TDC n |
| ASD mean age |
| ASD sd age |
| TDC mean age |
| TDC sd age |
| age range |
| Females n |
| control matching? |
| mean IQ ASD |
| mean IQ TDC |
| IQ > 70? |
| medicated? |
| Control group nature e.g. schools/hospital controls |
| Diagnosis |
| Diagnosis criteria |
| Scanner strength |
| pulse sequence |
| voxel size |
| voxel |
| TE |
| TR |
| shim |
| averages |
| points |
| fit residuals |
| other |
| software |
| quantification |
| sedation |
| Tissue correction |
| stat tests |
| multiple comparison corrections? |
| correlative findings. |

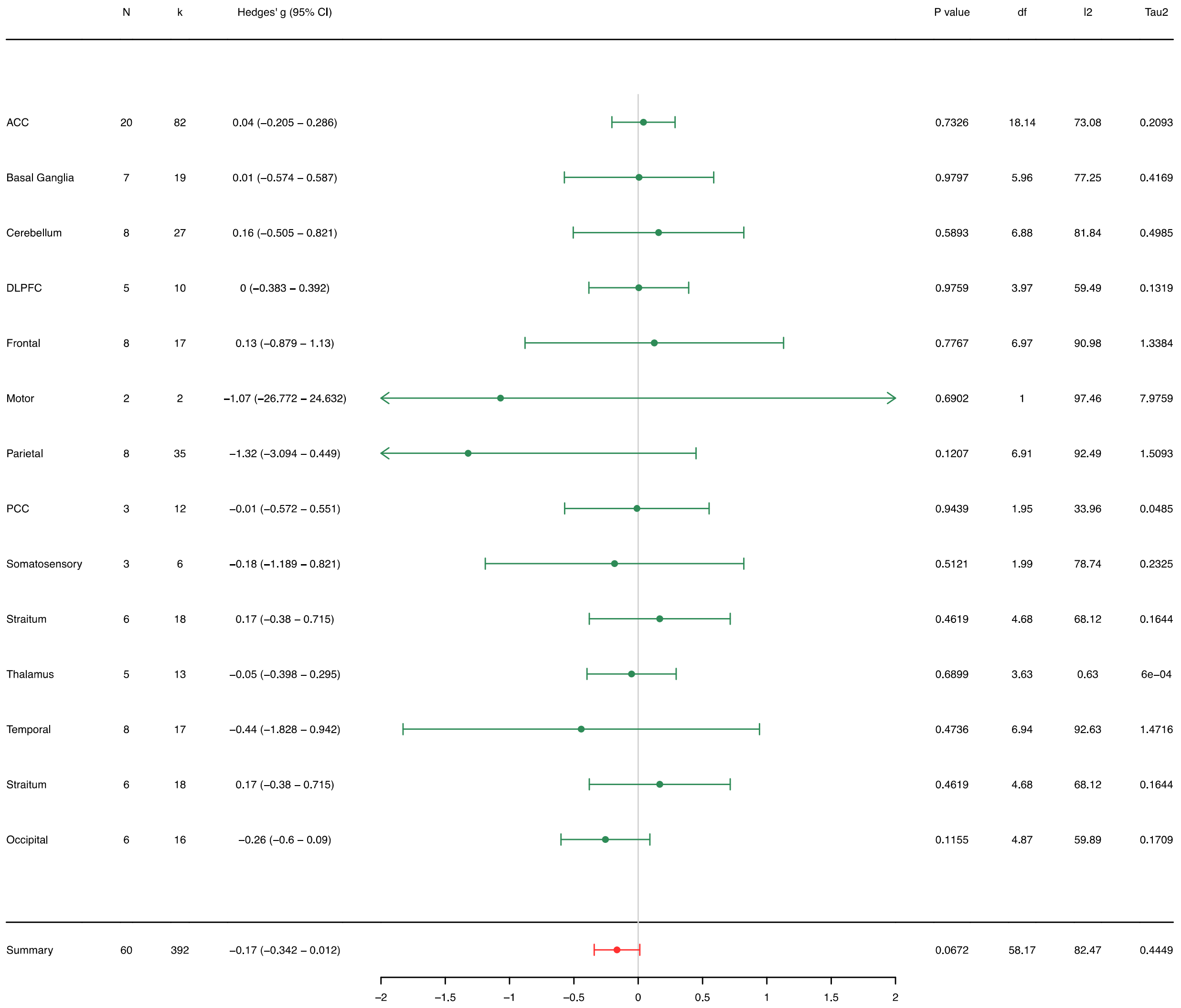

**Supplementary figure 1.** Summary forest plot for data grouped by brain region grouping 2. N: number of studies, k: number of observations, $I^2$: measure of between study heterogeneity, $Tau^2$: Variance in true effect sizes (another measure of between study heterogeneity). Hedge's g is reported in respect to autism. *Statistically significant at $p < 0.05$, and at $p < 0.01$ when the degrees of freedom < 4 for RVE t-tests. Note regions where n=1 are not shown on the forest plot. ACC = anterior cingulate cortex, DLPFC = dorsolateral prefrontal cortex, PCC = posterior cingulate cortex.

| Metabolite | N | k | Hedges' g (95% CI) | P value | df | I2 | Tau2 |
|---|---|---|---|---|---|---|---|
| **NAA** | | | | | | | |
| ACC | 14 | 16 | −0.22 (−0.428 – −0.005) | 0.0456* | 11.77 | 41.48 | 0.0527 |
| Basal Ganglia | 4 | 4 | −0.62 (−1.63 – 0.394) | 0.1459 | 2.95 | 65.7 | 0.2501 |
| Cerebellum | 7 | 8 | −0.34 (−0.823 – 0.139) | 0.1317 | 5.74 | 64.37 | 0.1755 |
| DLPFC | 6 | 7 | −0.11 (−0.546 – 0.326) | 0.5445 | 4.9 | 44.51 | 0.0787 |
| Frontal | 3 | 4 | −0.4 (−2.437 – 1.634) | 0.482 | 1.98 | 81.83 | 0.5251 |
| Medial temporal | 8 | 11 | −0.46 (−1.164 – 0.246) | 0.1676 | 6.94 | 81.16 | 0.5346 |
| Parietal | 6 | 10 | −0.34 (−0.514 – −0.156) | 0.0075* | 3.59 | 0 | 0 |
| PCC | 3 | 3 | −0.19 (−0.509 – 0.134) | 0.124 | 1.87 | 0 | 0 |
| Thalamus | 3 | 5 | −0.14 (−1.351 – 1.071) | 0.6645 | 1.97 | 56.49 | 0.1484 |
| Temporal | 2 | 4 | −0.24 (−12.988 – 12.517) | 0.8533 | 1 | 96.11 | 2.7864 |
| Summary | 35 | 85 | −0.32 (−0.544 – −0.099) | 0.0059* | 33.44 | 77.16 | 0.3254 |
| **Choline** | | | | | | | |
| ACC | 13 | 15 | 0.01 (−0.159 – 0.188) | 0.8565 | 9.06 | 9.08 | 0.0079 |
| Basal Ganglia | 3 | 3 | −0.22 (−2.607 – 2.169) | 0.7307 | 1.99 | 86.18 | 0.7982 |
| Cerebellum | 7 | 8 | 0.04 (−0.418 – 0.501) | 0.8299 | 5.62 | 59.91 | 0.1448 |
| DLPFC | 4 | 5 | −0.05 (−0.614 – 0.516) | 0.7956 | 2.89 | 24.46 | 0.0346 |
| Medial temporal | 8 | 11 | 0.23 (−0.384 – 0.847) | 0.4034 | 6.9 | 76.25 | 0.3868 |
| Parietal | 5 | 8 | −0.22 (−0.44 – −0.006) | 0.0462* | 3.66 | 0 | 0 |
| PCC | 3 | 3 | 0.29 (−0.867 – 1.455) | 0.3865 | 1.98 | 59.21 | 0.1253 |
| Summary | 27 | 63 | 0.07 (−0.064 – 0.213) | 0.2787 | 24.55 | 49.81 | 0.0937 |
| **Creatine** | | | | | | | |
| ACC | 9 | 11 | −0.25 (−0.625 – 0.125) | 0.1621 | 7.71 | 59.58 | 0.1373 |
| Basal Ganglia | 3 | 3 | −0.23 (−2.005 – 1.55) | 0.6338 | 1.98 | 74.02 | 0.355 |
| Cerebellum | 4 | 4 | −0.23 (−0.456 – −0.008) | 0.0462* | 2.65 | 0 | 0 |
| Medial temporal | 4 | 4 | 0.49 (−0.747 – 1.725) | 0.2958 | 2.98 | 74.04 | 0.4037 |
| Parietal | 3 | 5 | −0.11 (−0.564 – 0.352) | 0.4194 | 1.96 | 0 | 0 |
| Thalamus | 2 | 2 | 0.03 (−0.329 – 0.381) | 0.522 | 1 | 0 | 0 |
| Temporal | 2 | 3 | 0.28 (−10.437 – 10.998) | 0.7956 | 1 | 88.35 | 1.2591 |
| Summary | 27 | 59 | −0.15 (−0.395 – 0.09) | 0.2087 | 25.65 | 75.02 | 0.3109 |
| **Glx** | | | | | | | |
| ACC | 11 | 14 | −0.15 (−0.432 – 0.132) | 0.2607 | 9.46 | 60.08 | 0.1125 |
| Basal Ganglia | 4 | 4 | 0.05 (−1.516 – 1.613) | 0.9273 | 2.99 | 86.87 | 0.7737 |
| DLPFC | 3 | 3 | −0.2 (−0.576 – 0.166) | 0.1353 | 1.89 | 0 | 0 |
| Frontal | 2 | 2 | −0.54 (−16.441 – 15.363) | 0.741 | 1 | 95.21 | 2.9828 |
| Parietal | 5 | 8 | −1.02 (−3.255 – 1.211) | 0.2719 | 3.97 | 91.11 | 1.423 |
| PCC | 3 | 3 | −0.07 (−0.185 – 0.043) | 0.1115 | 1.87 | 0 | 0 |
| Thalamus | 4 | 6 | −0.15 (−0.471 – 0.177) | 0.2317 | 2.7 | 0 | 0 |
| Somatosensory | 2 | 2 | −0.17 (−6.965 – 6.627) | 0.805 | 1 | 88.58 | 0.5078 |
| Occipital | 4 | 6 | −0.44 (−1.704 – 0.826) | 0.3492 | 2.98 | 82.07 | 0.4426 |
| Summary | 29 | 61 | −0.08 (−0.316 – 0.151) | 0.4752 | 27.63 | 75.88 | 0.2875 |
| **Glu** | | | | | | | |
| ACC | 8 | 9 | 0.4 (−0.333 – 1.135) | 0.2349 | 6.65 | 84.04 | 0.3706 |
| Cerebellum | 3 | 3 | 0.58 (−1.779 – 2.948) | 0.3906 | 1.94 | 85.85 | 0.4743 |
| Straitum | 6 | 7 | 0.19 (−0.481 – 0.854) | 0.5001 | 4.76 | 74.69 | 0.2216 |
| Summary | 13 | 27 | 0.08 (−0.197 – 0.352) | 0.549 | 11.36 | 72.69 | 0.1997 |
| **GABA** | | | | | | | |
| ACC | 5 | 6 | −0.4 (−0.74 – −0.065) | 0.0336* | 2.58 | 0 | 0 |
| Cerebellum | 2 | 2 | −0.37 (−3.922 – 3.188) | 0.4146 | 1 | 54.62 | 0.092 |
| DLPFC | 3 | 3 | 0.31 (−1.155 – 1.766) | 0.4614 | 1.99 | 76 | 0.2756 |
| Frontal | 4 | 4 | −1.65 (−4.816 – 1.517) | 0.1953 | 2.98 | 92.39 | 2.1064 |
| Thalamus | 3 | 4 | −0.02 (−0.769 – 0.728) | 0.9149 | 1.95 | 13.45 | 0.0118 |
| Temporal | 5 | 5 | −0.52 (−3.187 – 2.145) | 0.6152 | 3.96 | 92.74 | 1.7889 |
| Somatosensory | 3 | 3 | 0.03 (−1.278 – 1.339) | 0.9296 | 1.99 | 77.4 | 0.2058 |
| Straitum | 2 | 2 | −0.1 (−0.88 – 0.683) | 0.3555 | 1 | 0 | 0 |
| Occipital | 6 | 8 | −0.09 (−0.455 – 0.267) | 0.5186 | 4.39 | 0 | 0 |
| Summary | 26 | 47 | −0.43 (−0.757 – −0.099) | 0.0129* | 24.6 | 82.99 | 0.4529 |
| **GSH** | | | | | | | |
| Limbic | 2 | 2 | −0.01 (−8.659 – 8.64) | 0.9908 | 1 | 90.11 | 0.8352 |
| Prefrontal | 2 | 2 | 0.3 (−4.904 – 5.502) | 0.5987 | 1 | 73.43 | 0.2463 |
| Summary | 3 | 6 | 0.1 (−1.275 – 1.483) | 0.7764 | 2 | 72.01 | 0.2543 |
| **mI** | | | | | | | |
| ACC | 8 | 10 | −0.21 (−0.636 – 0.216) | 0.2786 | 6.68 | 65.97 | 0.1553 |
| Basal Ganglia | 3 | 3 | −0.1 (−0.862 – 0.667) | 0.5941 | 1.72 | 0 | 0 |
| Cerebellum | 3 | 3 | 0.68 (−3.712 – 5.072) | 0.5707 | 1.98 | 91.96 | 1.3577 |
| Frontal | 3 | 3 | 0.85 (−0.527 – 2.237) | 0.1168 | 2 | 50.09 | 0.154 |
| Parietal | 3 | 6 | −2.22 (−10.224 – 5.792) | 0.3553 | 1.99 | 95.75 | 4.9329 |
| Summary | 20 | 44 | −0.04 (−0.352 – 0.271) | 0.7899 | 18.68 | 79.44 | 0.3947 |
| Summary | 60 | 392 | −0.17 (−0.342 – 0.012) | 0.0672 | 58.17 | 82.47 | 0.4449 |

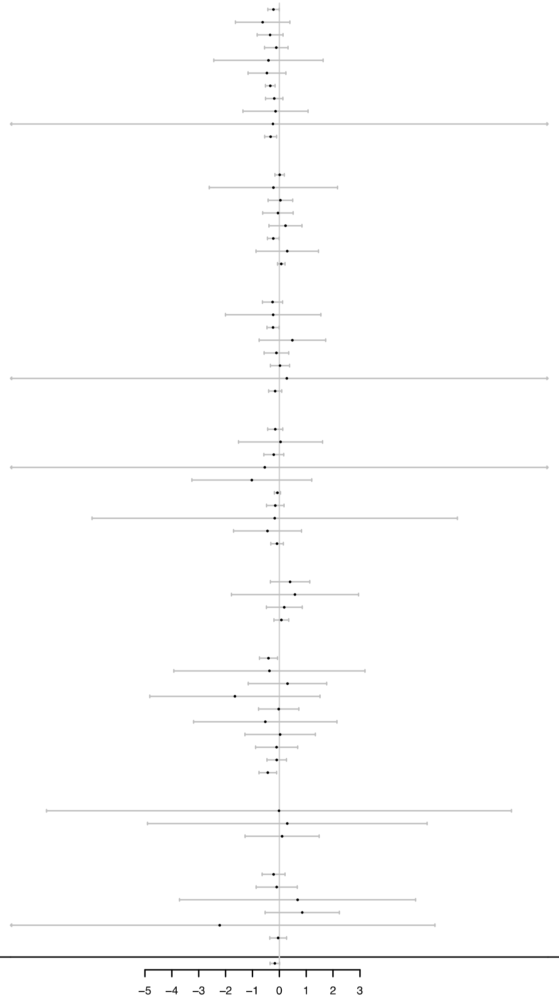

**Supplementary figure 2:** Summary Forest plot for data grouped by metabolite and brain region grouping 2. N: number of studies, k: number of observations, $I^2$: measure of between study heterogeneity, $Tau^2$: Variance in true effect sizes (another measure of between study heterogeneity). Hedge's g is reported in respect to autism. *Statistically significant at $p < 0.05$, and at $p < 0.01$ when the degrees of freedom < 4 for RVE t-tests. Note regions where n=1 are not shown on the forest plot.

| ACC | N | k | Hedges' g (95% CI) | P value | df | I2 | Tau2 |
|---|---|---|---|---|---|---|---|
| **Children** | | | | | | | |
| NAA | 7 | 8 | −0.11 (−0.395 – 0.172) | 0.3581 | 4.94 | 33.6 | 0.0324 |
| tCho | 6 | 7 | 0.02 (−0.326 – 0.373) | 0.8595 | 3.96 | 26.71 | 0.0269 |
| Creatine | 5 | 6 | −0.29 (−1.163 – 0.587) | 0.4099 | 3.92 | 75.85 | 0.3294 |
| Glx | 5 | 6 | 0 (−0.51 – 0.517) | 0.9847 | 3.55 | 52.4 | 0.0875 |
| Glu | 6 | 7 | 0.65 (−0.302 – 1.6) | 0.137 | 4.66 | 82.87 | 0.3683 |
| GABA | 4 | 8 | −0.5 (−0.669 – −0.322) | 0.0074* | 1.87 | 0 | 0 |
| MI | 5 | 6 | −0.23 (−0.906 – 0.437) | 0.3797 | 3.76 | 66.36 | 0.1591 |
| Summary | 12 | 44 | 0.2 (−0.222 – 0.631) | 0.3114 | 10.41 | 78.39 | 0.2731 |
| **Adults** | | | | | | | |
| NAA | 7 | 8 | −0.34 (−0.706 – 0.028) | 0.0647 | 5.82 | 41.4 | 0.0662 |
| tCho | 7 | 8 | 0 (−0.273 – 0.277) | 0.9878 | 5.5 | 5.24 | 0.0051 |
| Creatine | 4 | 5 | −0.27 (−0.654 – 0.117) | 0.1078 | 2.65 | 0 | 0 |
| Glx | 6 | 7 | −0.24 (−0.786 – 0.314) | 0.3185 | 4.93 | 73.55 | 0.2218 |
| Glu | 2 | 2 | −0.25 (−7.356 – 6.859) | 0.7339 | 1 | 89.46 | 0.56 |
| GABA | 1 | 2 | −0.21 (−0.637 – 0.221) | 0.34 | | | |
| MI | 3 | 4 | −0.18 (−1.593 – 1.229) | 0.6324 | 1.98 | 75.8 | 0.3063 |
| Summary | 8 | 36 | −0.15 (−0.417 – 0.114) | 0.2189 | 6.84 | 58.86 | 0.1235 |
| **Overall** | | | | | | | |
| NAA | 14 | 16 | −0.22 (−0.428 – −0.005) | 0.0456* | 11.77 | 41.48 | 0.0527 |
| tCho | 13 | 15 | 0.01 (−0.159 – 0.188) | 0.8565 | 9.06 | 9.08 | 0.0079 |
| Creatine | 9 | 11 | −0.25 (−0.625 – 0.125) | 0.1621 | 7.71 | 59.58 | 0.1373 |
| Glx | 11 | 13 | −0.11 (−0.425 – 0.197) | 0.4312 | 9.54 | 63.69 | 0.1309 |
| Glu | 8 | 9 | 0.4 (−0.333 – 1.135) | 0.2349 | 6.65 | 84.04 | 0.3706 |
| GABA | 5 | 5 | −0.43 (−0.674 – −0.18) | 0.0138* | 2.58 | 0 | 0 |
| MI | 8 | 10 | −0.21 (−0.636 – 0.216) | 0.2786 | 6.68 | 65.97 | 0.1553 |
| Summary | 20 | 80 | 0.05 (−0.199 – 0.294) | 0.6912 | 18.15 | 73.32 | 0.2118 |

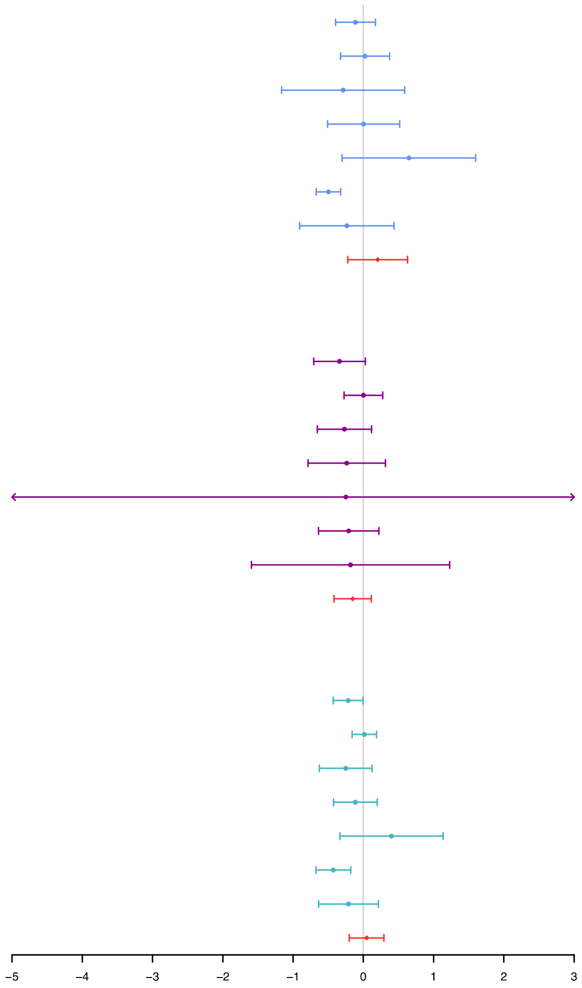

**Supplementary figure 3:** Summary Forest plot for data from the anterior cingulate cortex (ACC; brain grouping 2) grouped by metabolite and cohort age. N: number of studies, k: number of observations, $I^2$: measure of between study heterogeneity, $Tau^2$: Variance in true effect sizes (another measure of between study heterogeneity). Hedge's g is reported in respect to autism. *Statistically significant at $p < 0.05$, and at $p < 0.01$ when the degrees of freedom < 4 for RVE t-tests. Note regions where n=1 are not shown on the forest plot.

| Demographics/Methods GABA | N | k | Hedges' g (95% CI) | P value | df | I2 | Tau2 |
|---|---|---|---|---|---|---|---|
| **50% female** | | | | | | | |
| Y | 5 | 7 | −0.03 (−0.329 – 0.26) | 0.7599 | 3.8 | 0 | 0 |
| N | 19 | 33 | −0.54 (−1.002 – −0.082) | 0.0234* | 17.71 | 86.09 | 0.5392 |
| ? | 3 | 5 | −0.39 (−1.283 – 0.504) | 0.1968 | 1.94 | 60.63 | 0.1872 |
| **1:3 female:male** | | | | | | | |
| Y | 7 | 14 | −0.39 (−1.202 – 0.429) | 0.2891 | 5.92 | 83.26 | 0.5645 |
| N | 16 | 26 | −0.48 (−0.951 – −0.019) | 0.0424* | 14.74 | 84.4 | 0.4603 |
| ? | 3 | 5 | −0.39 (−1.283 – 0.504) | 0.1968 | 1.94 | 60.63 | 0.1872 |
| **IQ > 70 only** | | | | | | | |
| Y | 19 | 32 | −0.27 (−0.559 – 0.022) | 0.0684 | 17.71 | 74.26 | 0.2568 |
| N | 3 | 6 | −1.16 (−4.358 – 2.045) | 0.258 | 1.98 | 91.09 | 1.1963 |
| ? | 4 | 7 | −0.91 (−3.055 – 1.229) | 0.2678 | 2.99 | 91.22 | 1.8694 |
| **Participants on psychotropic medications** | | | | | | | |
| Y | 18 | 29 | −0.45 (−0.844 – −0.051) | 0.0293* | 16.71 | 80.98 | 0.3975 |
| N | 3 | 5 | 0.01 (−0.401 – 0.421) | 0.9198 | 1.8 | 0 | 0 |
| ? | 5 | 11 | −0.78 (−2.171 – 0.607) | 0.1925 | 3.96 | 90.12 | 1.0273 |
| **Use of sedation** | | | | | | | |
| Y | 4 | 7 | −0.52 (−0.802 – −0.248) | 0.0113* | 2.52 | 39.3 | 0.0614 |
| N | 22 | 38 | −0.44 (−0.831 – −0.041) | 0.0322* | 20.69 | 83.82 | 0.5033 |
| **Reported diagnosis** | | | | | | | |
| ASD | 24 | 43 | −0.41 (−0.764 – −0.048) | 0.0271* | | | |
| AD | 1 | 2 | −2.92 (−26.2 – 20.77) | 0.3613 | | | |
| AS | 1 | 2 | −0.33 (−3.99 – 3.995) | 0.461 | | | |
| **Quantification method** | | | | | | | |
| i.u (water referenced) | 19 | 33 | −0.11 (−0.249 – 0.02) | 0.0905 | 17.1 | 37.4 | 0.0521 |
| /Cr (creatine referenced) | 7 | 12 | −1.62 (−2.97 – −0.27) | 0.0261* | 5.97 | 92.39 | 1.637 |
| **Tissue correction** | | | | | | | |
| Y | 15 | 26 | −0.07 (−0.224 – 0.078) | 0.3176 | 13.25 | 30.39 | 0.0353 |
| N | 4 | 7 | −0.34 (−0.829 – 0.154) | 0.1161 | 2.95 | 56.54 | 0.1685 |
| **Scanner field strength** | | | | | | | |
| 1.5T | 1 | 3 | −2.92 (−26.62 – 20.77) | 0.3613 | | | |
| 3T | 25 | 45 | −0.4 (−0.735 – −0.058) | 0.9843 | | | |
| **MRS pulse sequence** | | | | | | | |
| MEGA−PRESS | 17 | 21 | −0.52 (−0.922 – −0.123) | 0.0096* | | | |
| HERMES | 1 | 3 | −0.14 (−1.638 – 1.362) | 0.459 | | | |
| MEGA−SPECIAL | 1 | 1 | −0.01 (−3.87 – 2.822) | 0.9843 | | | |
| STEAM | 1 | 1 | −0.78 (−10.99 – 9.433) | 0.5099 | | | |
| Summary | 26 | 45 | −0.44 (−0.761 – −0.121) | 0.009* | 24.56 | 81.98 | 0.4191 |

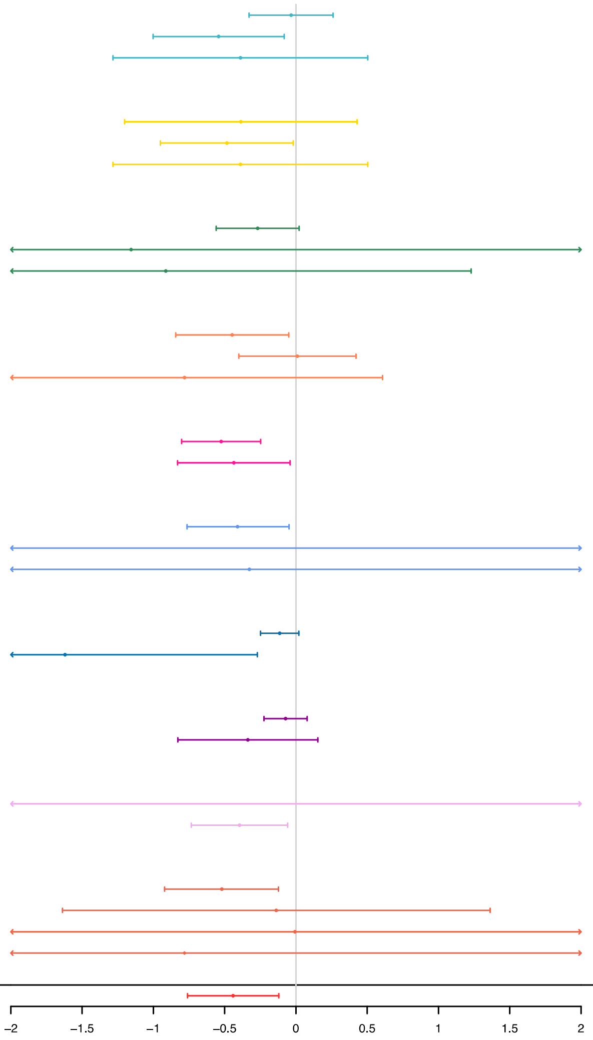

**Supplementary figure 4**. Forest plot summary of GABA data grouped by demographic factors and MRS acquisition parameters. N: number of studies, k: number of observations, % children: percentage of studies observing children only, $I^2$: measure of between study heterogeneity, $Tau^2$: Variance in true effect sizes (another measure of between study heterogeneity). Hedges' g is reported in respect to autism. *Statistically significant at $p < 0.05$, and at $p < 0.01$ when the degrees of freedom < 4.

| Demographics/Methods: NAA | N | k | Hedges' g (95% CI) | P value | df | I2 | Tau2 |
|---|---|---|---|---|---|---|---|
| **50% female** | | | | | | | |
| Y | 2 | 5 | −0.55 (−5.843 – 4.752) | 0.4152 | 1 | 86.19 | 0.9268 |
| N | 30 | 71 | −0.24 (−0.417 – −0.054) | 0.013* | 28.36 | 68.96 | 0.2074 |
| ? | 4 | 9 | −1.29 (−4.414 – 1.841) | 0.2813 | 2.99 | 93.49 | 2.182 |
| **1:3 female:male** | | | | | | | |
| Y | 4 | 12 | −0.57 (−1.68 – 0.539) | 0.1998 | 2.98 | 81.47 | 0.4953 |
| N | 25 | 60 | −0.21 (−0.414 – −0.005) | 0.0452* | 23.43 | 69.19 | 0.209 |
| ? | 7 | 13 | −0.71 (−1.98 – 0.554) | 0.2171 | 5.95 | 88.67 | 0.9521 |
| **IQ > 70 only** | | | | | | | |
| Y | 22 | 49 | −0.23 (−0.594 – 0.128) | 0.194 | 20.78 | 80.15 | 0.4687 |
| N | 5 | 15 | −0.37 (−0.86 – 0.116) | 0.1012 | 3.88 | 73.68 | 0.1932 |
| ? | 8 | 21 | −0.54 (−0.83 – −0.25) | 0.0033* | 6.75 | 62.32 | 0.1521 |
| **Participants on psychotropic medications** | | | | | | | |
| Y | 10 | 18 | −0.45 (−1.339 – 0.436) | 0.2796 | 8.94 | 87.48 | 0.892 |
| N | 12 | 35 | −0.17 (−0.462 – 0.124) | 0.231 | 10.82 | 68.29 | 0.2576 |
| ? | 13 | 32 | −0.39 (−0.646 – −0.141) | 0.0054* | 11.61 | 68.36 | 0.1577 |
| **Use of sedation** | | | | | | | |
| Y | 7 | 13 | −0.15 (−0.588 – 0.288) | 0.4289 | 5.66 | 64.9 | 0.1425 |
| N | 28 | 72 | −0.38 (−0.655 – −0.098) | 0.01* | 26.68 | 79.22 | 0.4042 |
| **Reported diagnosis** | | | | | | | |
| ASD | 24 | 57 | −0.31 (−0.614 – −0.015) | 0.0406* | 22.59 | 78.6 | 0.3442 |
| AD | 7 | 20 | −0.61 (−0.97 – −0.251) | 0.0061* | 5.87 | 70.53 | 0.2387 |
| ASP | 4 | 8 | 0.16 (−0.492 – 0.822) | 0.4822 | 2.98 | 67.32 | 0.2873 |
| **Quantification method** | | | | | | | |
| /Cr (creatine referenced) | 27 | 65 | −0.29 (−0.558 – −0.031) | 0.0298* | 25.61 | 77.18 | 0.3402 |
| creatine | 8 | 20 | −0.42 (−0.957 – 0.117) | 0.1069 | 6.88 | 79.46 | 0.3596 |
| **Tissue correction** | | | | | | | |
| Y | 14 | 25 | −0.2 (−0.433 – 0.04) | 0.095 | 12.61 | 55.73 | 0.1268 |
| N | 11 | 30 | −0.41 (−1.152 – 0.339) | 0.2519 | 9.91 | 85.9 | 0.7086 |
| **Scanner field strength** | | | | | | | |
| 1.5T | 19 | 51 | −0.47 (−0.906 – −0.037) | 0.0351* | 17.83 | 84.09 | 0.5992 |
| 3T | 15 | 32 | −0.16 (−0.373 – 0.054) | 0.13 | 13.41 | 52.39 | 0.0938 |
| **MRS pulse sequence** | | | | | | | |
| PRESS | 45 | 254 | 0.08 (−0.057 – 0.212) | 0.252 | 43.03 | 70.94 | 0.2314 |
| MEGA−PRESS | 21 | 34 | −0.52 (−0.922 – −0.123) | 0.013* | 19.64 | 83.78 | 0.4626 |
| HERMES | 2 | 8 | −0.42 (−6.71 – 5.868) | 0.5513 | 1 | 83.82 | 0.5255 |
| STEAM | 14 | 74 | −0.37 (−0.709 – −0.039) | 0.0315* | 12.76 | 78.66 | 0.3362 |
| PROBE−P | 1 | 1 | 0.77 (−5.252 – 6.79) | 0.3512 | | | |
| MEGA−SPECIAL | 1 | 1 | 0.25 (−0.596 – 1.095) | 0.1659 | | | |
| Summary | 35 | 85 | −0.32 (−0.544 – −0.099) | 0.0059* | 33.44 | 77.16 | 0.3254 |

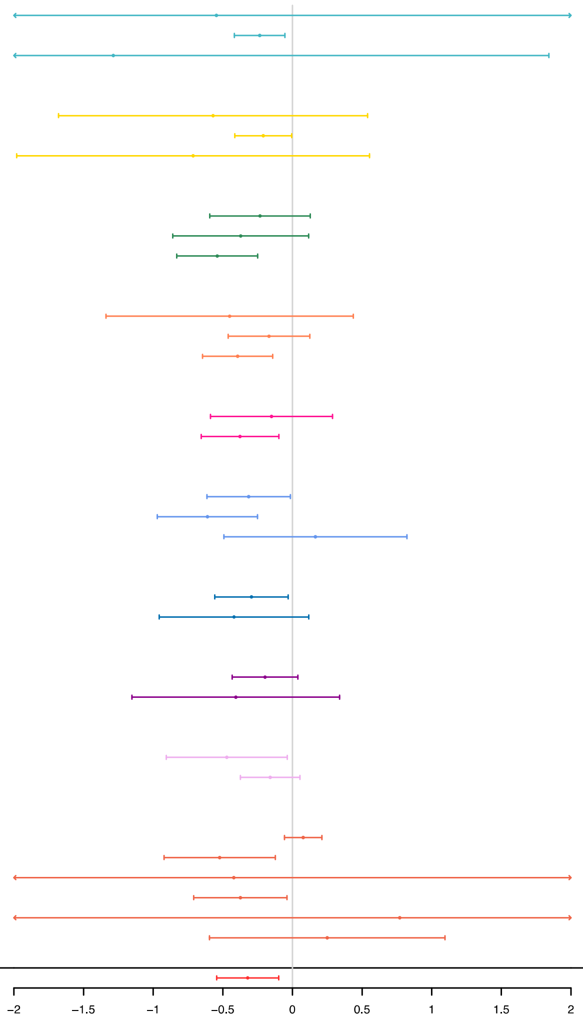

**Supplementary figure 5**. Forest plot summary of NAA data grouped by demographic factors and MRS acquisition parameters. N: number of studies, k: number of observations, % children: percentage of studies observing children only, $I^2$: measure of between study heterogeneity, $Tau^2$: Variance in true effect sizes (another measure of between study heterogeneity). **Hedges' g is reported in respect to autism.** *Statistically significant at $p < 0.05$, and at $p < 0.01$ when the degrees of freedom < 4.

**Supplementary table 2**: Brain region grouping 1.

Brain regions were grouped into broader regions of interest for the meta-analysis.

| Region | Brain regions included |
|---|---|
| Limbic | thalamus, basal ganglia, lenticular nuclei, striatum, medial temporal lobe, cingulate, caudate, putamen, hippocampus, pregenual anterior cingulate. |
| Frontal | frontal lobe, inferior frontal gyrus, frontal eye fields, motor cortex. |
| Temporal | temporal lobe, temporal gyrus, superior temporal sulcus, auditory cortex. |
| Cerebellum | cerebellum, cerebellar vermis |
| Prefrontal | medial prefrontal, dorsolateral prefrontal cortex, orbitofrontal cortex, dorsomedial prefrontal cortex. |
| Insular | Insular |
| Occipital | occipital lobe, occiput, visual cortex, MT+ area. |

| | |
|---|---|
| Parietal | parietal lobe, temporal parietal lobe, intraparietal sulcus, posterior parietal. |
| White matter (WM) | WM, callosum |
| Grey matter (GM) | WM |

**Supplementary table 3:** Brain region grouping 2.

Brain regions were grouped into more specific brain region groupings that were used to explore significant results with greater spatial resolution. Study number was considerably reduced when data was grouped this way and as such brain region grouping 1 was preferred for the main meta-analysis.

| Region | brain regions included |
|---|---|
| Thalamus | thalamus |
| Basal Ganglia | basal ganglia, caudate, putamen |
| Striatum | striatum, lenticular nuclei |
| Medial temporal lobe | hypothalamus-amygdala area, hippocampus, medial temporal lobe. |
| Anterior cingulate cortex (ACC) | pregenual anterior cingulate (ACC) |
| Dorsolateral prefrontal cortex (DLPFC) | dorsolateral prefrontal cortex (DLPFC) |
| Posterior cingulate cortex (PCC) | posterior cingulate cortex (PCC) |

| | |
|---|---|
| Frontal | frontal lobe, Inferior frontal gyrus, frontal eye fields. |
| Motor cortex | motor cortex, sensory motor area. |
| Temporal | temporal lobe, temporal gyrus, superior temporal sulcus, auditory cortex. |
| Cerebellum | cerebellum, cerebellar vermis |
| Somatosensory cortex | somatosensory cortex |
| Prefrontal | medial prefrontal, orbitofrontal cortex. |
| Insular | Insular |
| Occipital | occipital lobe, occiput, visual cortex, MT+ area. |
| Parietal | parietal lobe, temporal parietal lobe, intraparietal sulcus, posterior parietal. |
| White matter (WM) | WM, callosum |
| Grey matter (GM) | WM |
